# NeuroMorph: A Unified Morphological Reference Space for Cross-Disease Brain Profiling

**DOI:** 10.64898/2026.08.13.26359403

**Authors:** Connor Dalby, Austin Dibble, Sergio Benini, Damiano Ferrari, Donald M. Lyall, Monika Harvey, Terry Quinn, Lars Muckli, Alessio Fracasso, Michele Svanera, Alzheimer’s Disease Neuroimaging Initiative, Frontotemporal Lobar Degeneration Neuroimaging Initiative

## Abstract

Structural MRI is routinely acquired in clinical practice, yet quantitative morphometry has had limited impact on clinical decision-making. Overlapping symptoms, trajectories and comorbidities remain difficult to interpret within disease-specific frameworks, leaving it unclear how individual patients relate to the broader organization of brain disease. Here, we construct a cross-disease morphological reference space from 110,591 T1w MRI scans of 78,794 participants, spanning four disease families, 19 diagnoses, and seven subtypes. To construct this space, we developed NeuroMorph, an AI framework deriving thirteen interpretable morphological descriptors and individual normative deviation profiles. The reference space reveals shared and distinct morphological signatures that distinguish conditions within a hierarchy of disease families, diagnoses, subtypes and individual profiles. It identifies overlapping and comorbid morphological profiles and captures longitudinal deviations that precede clinical diagnosis and track progression. Together, these findings establish a unified framework for mapping brain disease organization and positioning individual patients within its morphological landscape.

## Main

Structural MRI (sMRI) is routinely acquired in clinical practice and provides a scalable window into neuroanatomical integrity. Yet, despite decades of neuroimaging research, quantitative morphometry derived from sMRI has had limited impact on individual-level decision-making in neurology and psychiatry [1, 2]. Although diagnostic labels remain essential for patient care and provide the primary framework for clinical decision-making, many neurological and psychiatric conditions share symptoms, risk factors and trajectories, or co-occur in ways that are difficult to interpret within disease-specific frameworks alone [3]. Whether these clinical overlaps correspond to shared or distinct patterns of brain morphology remains difficult to quantify, preventing individual patients from being interpreted within the broader organization of brain disease.

Over the past two decades, sMRI studies have characterized brain morphology across the lifespan, within population cohorts, and in a wide range of neurological and psychiatric conditions [4–6]. Traditional voxel-based and surface-based morphometric studies have established important disease-associated patterns of atrophy and cortical change, but have relied primarily on group-level comparisons and have often examined disorders in isolation [7, 8]. Normative modeling has advanced the field by shifting the focus from average case-control differences to individual deviations from age- and sex-appropriate reference distributions [9, 10]. However, most applications remain focused on selected disorders, limited morphological features or specific clinical tasks such as classification and prognosis, leaving it unclear how individual morphological deviations relate to one another across disorders [11].

The remaining challenge is therefore not only to determine whether an individual brain differs from expectation, but also to place that difference within a shared morphology space [12, 13]. Such a reference space would allow brain disorders to be compared directly, revealing how conditions cluster into broader disease families, where clinically overlapping disorders diverge, and whether comorbid presentations occupy intermediate or convergent morphological deviation profiles. It would also provide an anatomical context for interpreting individual patients according to their morphological similarity across disorders rather than within isolated diagnostic categories. By linking population-level structure with patient-level variation, this morphology space could characterize how individual patients relate to the broader organization of disease, with relevance for stratification, comorbidity and longitudinal monitoring [9, 14, 15].

Here, we establish a scalable cross-disease morphological reference space from structural MRI. This space maps brain morphology across a hierarchy spanning healthy populations, disease families, diagnoses, subtypes and patient-level morphological deviation profiles. To enable this, we developed NeuroMorph, an AI-based morphometric framework for rapid and robust estimation of brain morphology from T1-weighted scans. Applied to 110,591 T1-weighted MRI scans across 19 neurological and psychiatric conditions, NeuroMorph derives compact morphological fingerprints and corresponding normative morphological deviation profiles suitable for large-scale cross-disease analysis.

We show how the morphological reference space is organized across neurological and psychiatric disease, supporting applications including differential characterization, comorbidity mapping, and longitudinal tracking. At the individual level, the space contextualizes a subject’s morphological deviation profile, identifies patients whose anatomical profiles resemble related or overlapping conditions, and tracks whether brain structure follows an expected or atypical trajectory over time, providing a basis for stratification, monitoring and prognostic assessment. Together, these findings establish a cross-disease morphological reference space that provides a foundation for understanding how brain disease is organized, and a means of situating individual patients within its broader landscape. NeuroMorph code, model, and documentation are available on this GitHub repository (https://github.com/rockNroll87q/NeuroMorph)

## Results

### Defining The Morphology Reference space

To establish a hierarchical morphology reference space, we first derived compact individual representations of brain structure from routine T1-weighted MRI using NeuroMorph (Method 1). NeuroMorph integrates two components, LOD Brain+ (Method 1a) and DeepThickness (Method 1b), which generate segmentation masks and cortical surface reconstructions, respectively. Both methods were benchmarked against FreeSurfer for speed, accuracy and expert preference (Method 1c). For each scan, the framework extracts 13 complementary morphological descriptors spanning cortical thickness, surface geometry, and regional tissue volumes, generating an individual morphological *fingerprint*. Normative modeling subsequently transformed these fingerprints into morphological deviation *profiles*, representing each individual’s z-scored departure from age- and sex-matched healthy controls (Fig. 1a, Method 2). Morphological fingerprints therefore describe brain structure, whereas deviation profiles quantify departures from healthy normative ageing.

**Fig. 1:**
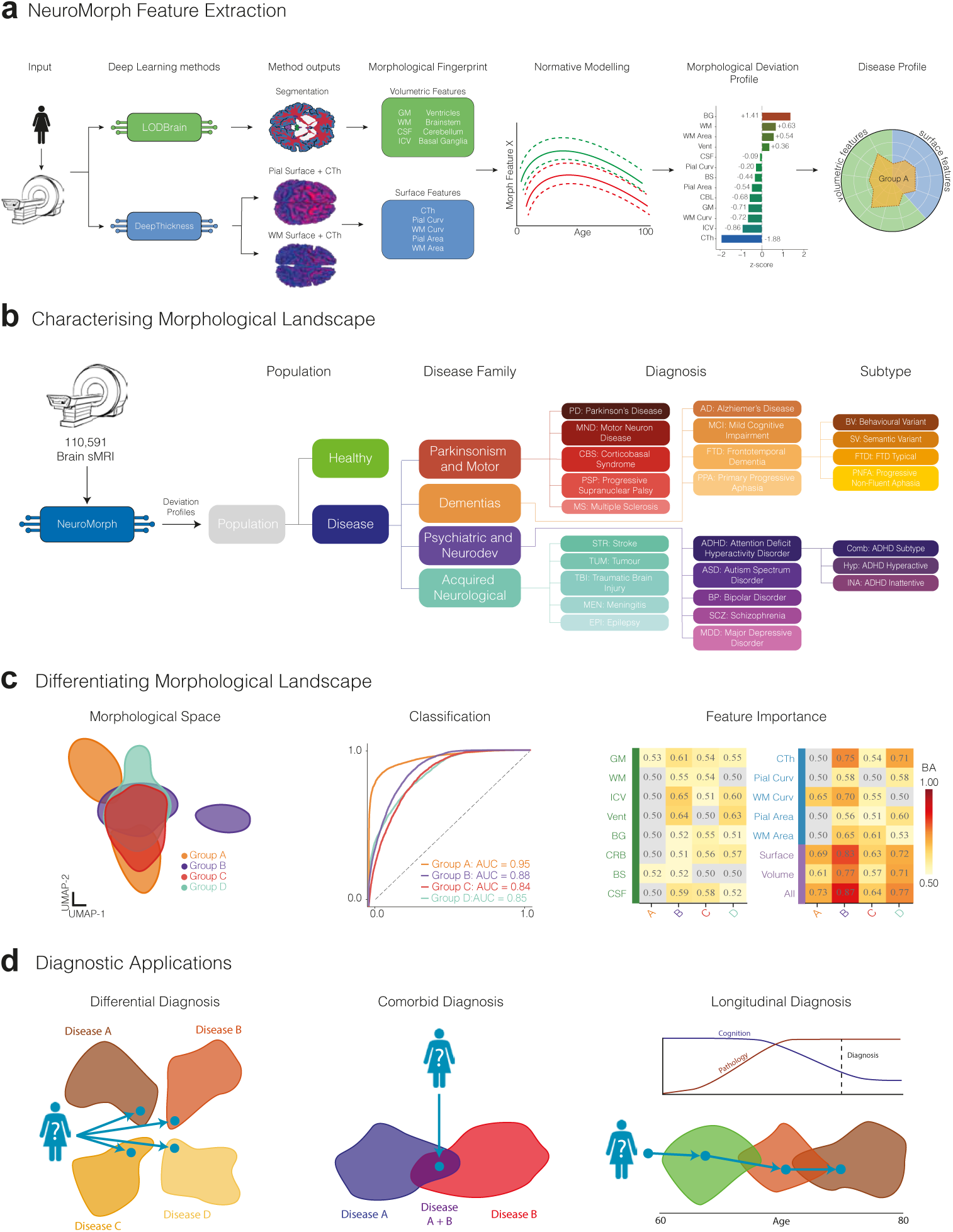
Study overview. **a,** The NeuroMorph framework extracts 13 morphological features from native-space T1-weighted sMRI. NeuroMorph integrates two deep learning models, LOD Brain+ and DeepThickness, which generate segmentation masks and cortical surface reconstructions to derive eight volumetric and five surface-based features respectively. **b,** The disease hierarchy comprises 110,591 sMRI scans from 78,794 participants, spanning four levels of resolution from population, through disease family and diagnosis, to subtype. **c,** The analysis workflow applied at each level of the hierarchy: defining the shared morphological space, classifying conditions and quantifying f_4_eature importance. **d,** Downstream applications of the shared morphological space at the group and individual levels, comprising, from left to right, differential, comorbid and longitudinal diagnosis.

We applied this framework to 110,591 sMRI scans spanning healthy controls, four disease families, 19 diagnoses, and seven disease subtypes (Fig. 1b). At each level of this hierarchy, morphological deviation profiles defined a morphology reference space in which we asked whether morphology distinguished conditions at the same level, which anatomical descriptors drove this separation, and how individual patients were positioned relative to one another (Fig. 1c,d). This design enabled a unified hierarchical analysis spanning health-disease separation, disease-family organization, diagnosis-level stratification and individual patient positioning within a common morphology reference space.

### The Morphology Reference Space Stratifies Across a Disease Hierarchy

We first investigated whether the morphology reference space preserves the hierarchical organization of brain disorders across four levels, descending from the whole population to disease family, diagnosis and subtype. At each level, we examined the following three properties: how conditions were organized within the reference space (i.e., organization), whether this organization supported quantitative discrimination between groups (i.e., differentiation), and which morphological descriptors defined their separation (i.e., interpretation). Throughout, we tracked the Dementias family as a representative example (Fig. 2, col. I). This family contains labels down to the subtype level, with known overlap in clinical presentations between groups at each level of the hierarchy. The same analysis across the remaining disease families is shown in Extended Data Fig. 1.

**Fig. 2:**
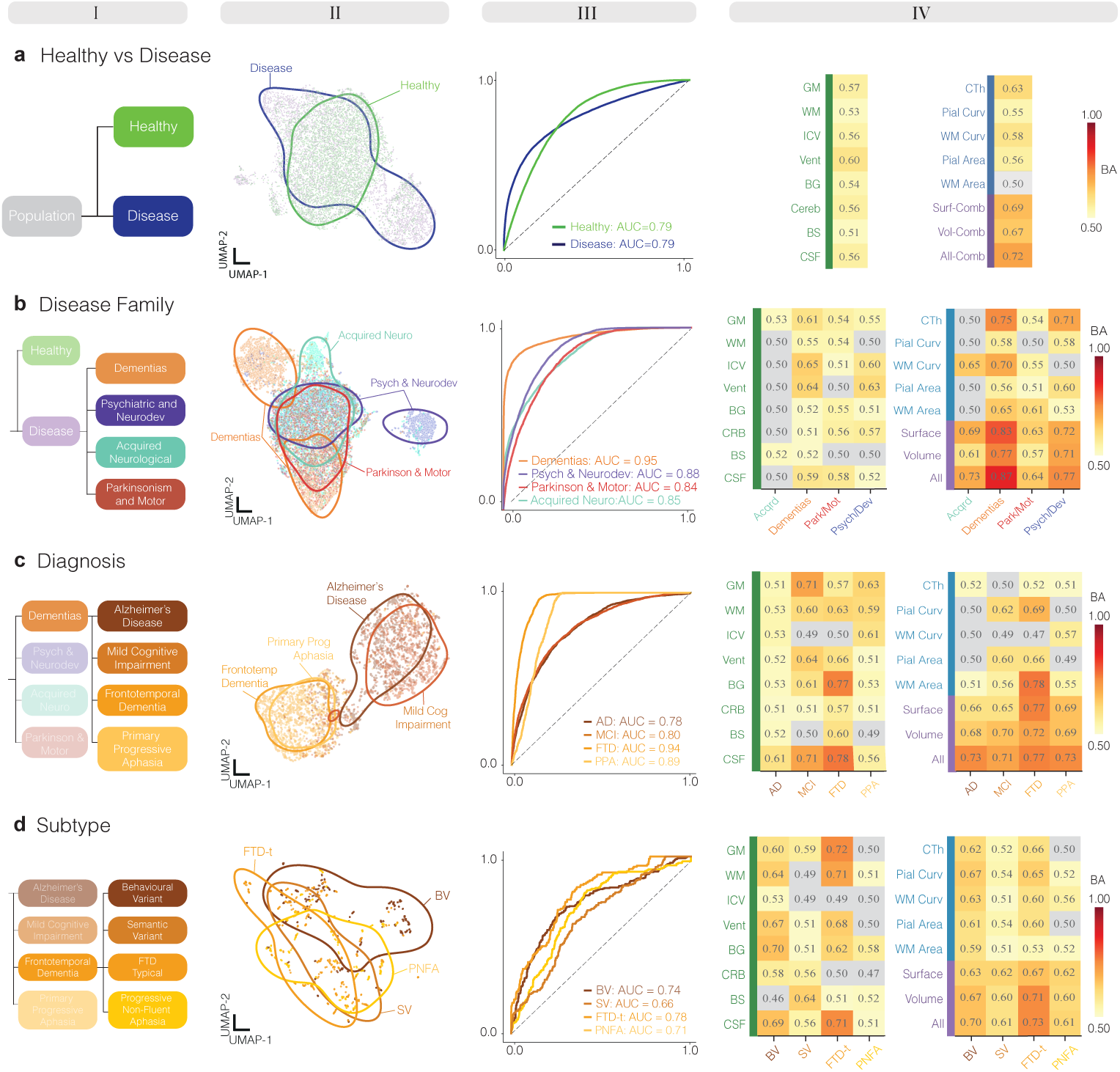
Morphological Stratification Across a Disease Hierarchy. *Column I:* Position within the disease hierarchy, indicating which conditions are included at each level of analysis. *Column II:* UMAP projection of the shared morphological deviation space, in which each point represents a single individual’s morphological deviation profile across the 13 NeuroMorph descriptors. These projections are used for visualization only; quantitative discrimination is assessed in III and IV. *Column III:* Receiver operating characteristic (ROC) curves for each condition at the current hierarchical level, derived from one-versus-rest binary (**a**) or multiclass (**b-d**) classification. *Column IV:* Descriptor-set performance heatmap in which each entry represents the balanced accuracy achieved by a given descriptor set when differentiating each condition from all others at the current level. Row labels in green and blue correspond to volumetric and surface-based descriptors evaluated in isolation, respectively, while purple labels denote combined descriptor sets incorporating all volumetric, surface-based, or morphological descriptors. Balanced accuracy values approaching 1.0 indicate strong discrimination (red), while values approaching 0.5 reflect chance-level performance (white).

**Extended Data Fig.1:**
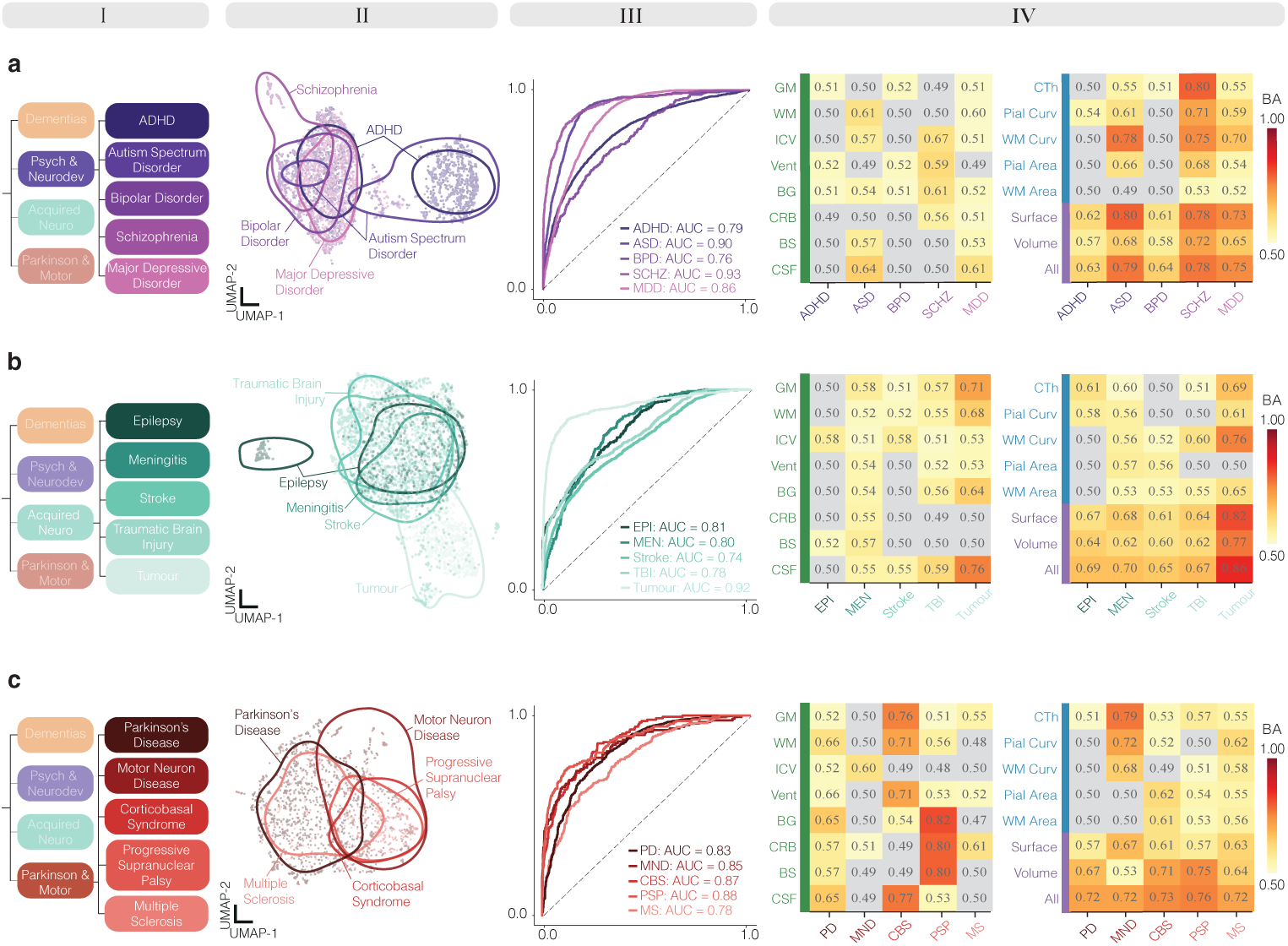
Diagnosis stratification across three disease families. Each row represents the differentiation results, at the diagnosis level, within a specific disease family. *Column I:* Position within the disease hierarchy, indicating which conditions are included at each level of analysis. *Column II:* UMAP projection of the shared morphological feature space, in which each point represents a single individual. *Column III:* ROC curves for each condition at the current hierarchical level, derived from multiclass and binary (*≤*2 conditions) classification. *Column IV:* Feature profile importance heat map in which each entry represents the balanced accuracy achieved by a given feature profile set when differentiating each condition from all others at the current level. Balanced accuracy values approaching 1.0 indicate strong discrimination (red), while values approaching 0.5 reflect chance-level performance (white).

The resulting reference space was visualized by projecting 13D morphological deviation profiles at each hierarchical level into a 2D common UMAP representation (Fig.2, col. II, Method 3). At the population and disease family levels, groups shared a large common region, reflecting variation present across that level, while several groups also occupied peripheral pockets distinct from this substrate. Separation sharpened as the hierarchy narrowed. At the diagnosis level, clinically similar conditions, such as Alzheimer’s Disease (AD) and mild cognitive impairment (MCI), or frontotemporal dementia (FTD) and primary progressive aphasia (PPA), overlapped closely, yet the two pairs remained clearly separated. At the subtype level, FTD divided further into its behavioral, semantic and non-fluent variants, although with more pronounced overlap than at broader levels. These observations suggest that neighbouring regions of the reference space reflect biological similarity across disorders while preserving hierarchical disease structure. The morphological profile of the complete hierarchy (all levels) is also visualized via UMAP projections in Supplementary Figs 1 and 2, and radar plots in Supplementary Figs. 3-5.

We next asked whether this organization retained sufficient information to distinguish conditions quantitatively despite their partial overlap in the UMAP embedding. To evaluate the separability of the reference space, we trained random forest classifiers using the complete 13-feature morphological deviation profiles, which differentiated groups at every level of the hierarchy (Fig. 2, col. III, Method 4). Binary classification (Method 4a) distinguished healthy from disease individuals (AUC = 0.79), and multiclass classification (Method 4b) resolved even the overlapping pairs, separating FTD from PPA and AD from MCI. Performance peaked at the disease family level and declined only marginally as distinctions became increasingly fine-grained toward diagnosis and subtype levels.

Finally, we investigated which morphological descriptors defined the organization of the reference space and supported its discriminative power (Fig. 2, col. IV, Method 4). Individual descriptors, including cortical thickness, gray matter and white matter volumes, often reached high balanced accuracy on their own. However, combined descriptor sets (whether the volumetric set, the surface set, or all 13 together) consistently produced the strongest differentiation at every level, exceeding the accuracy of any single descriptor used alone. Balanced accuracy heatmaps across all hierarchy levels are shown in Supplementary Fig. 6. Centile brain charts for each morphological descriptor in the healthy and disease groups are also shown in Supplementary Fig. 7 .

Together, these results demonstrate that the morphology reference space preserves the hierarchical organization of brain disorders by integrating multiple morphological descriptors that support robust discrimination from broad disease families to clinically overlapping subtypes.

### The Morphology Reference Space Supports Differential Characterization

We next asked whether the morphology reference space could support differential characterization among conditions with overlapping clinical and morphological presentations. This analysis was performed across the hierarchy; here, we focus on the Parkinsonism and Motor family as a representative case study (Fig. 3), with contrasts for other disease families reported in Extended Data Fig. 2. Parkinson’s disease (PD), progressive supranuclear palsy (PSP) and corticobasal syndrome (CBS) provide an informative example because of their overlap in clinical presentations [16, 17]. To evaluate how these clinically related disorders were represented within the reference space, we examined group-level deviation profiles, their position within the shared morphological space, age-dependent trajectories of discriminative descriptors and quantitative classification performance (Fig. 3a,b). Finally, we asked whether this representation could support interpretation of an individual patient with PD (Fig. 3c).

**Fig. 3:**
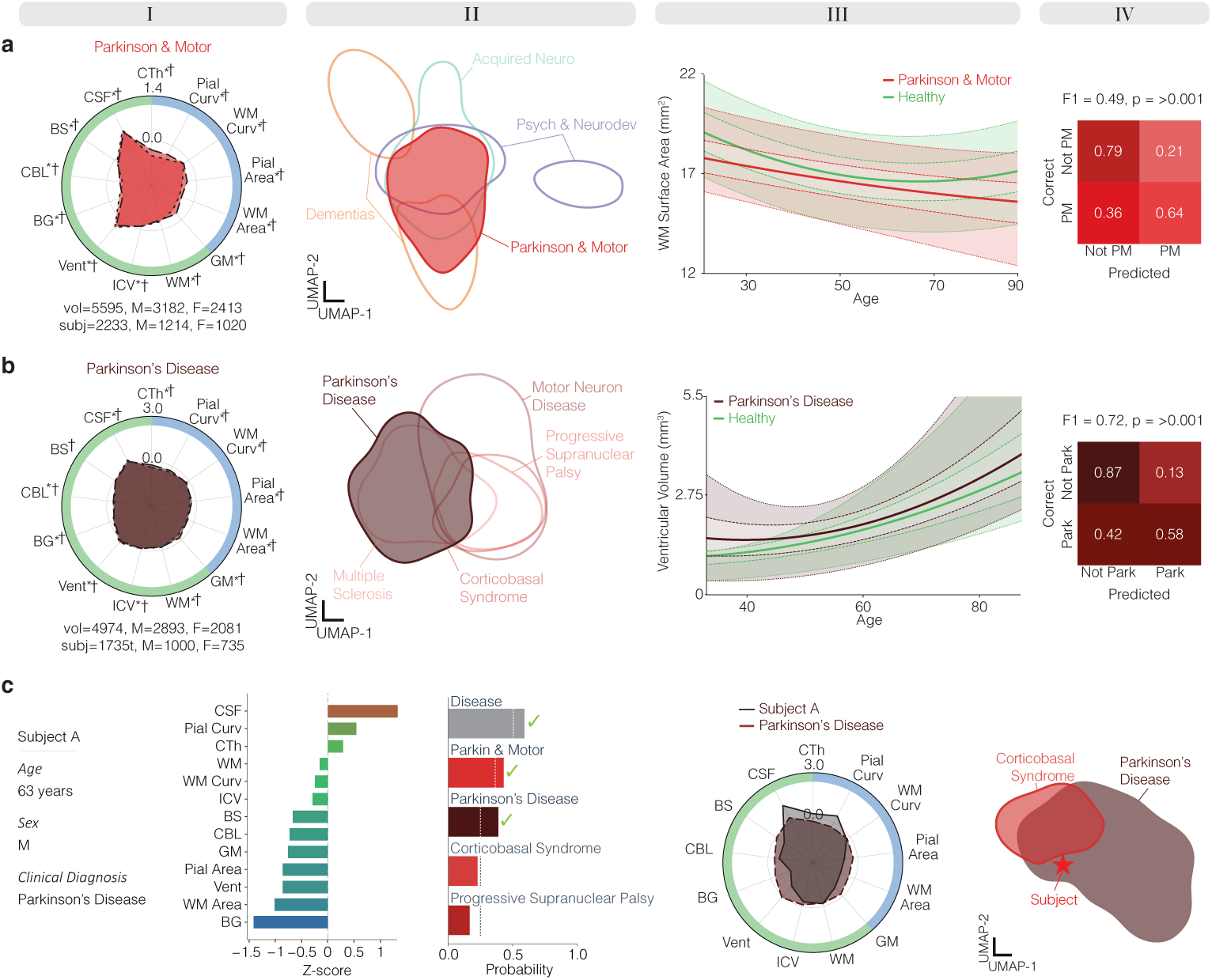
Morphological differentiation across a disease hierarchy. **a-b,** *Column I:* Morphological feature profile of the target condition at the family (**a**) and diagnosis (**b**) level, represented as a radar plot. The centre circle represents the age and sex controlled healthy cohort. Significant differences between male and female cohorts are denoted by * and *†*, respectively. *Column II:* UMAP projection of the shared morphological feature space across conditions at the family (**a**) and diagnosis (**b**) level. *Column III:* Normative trajectories for the target condition relative to the healthy group across the age range of the target cohort. Normative centiles are represented as follows: 0.5 (median) as a bold line, 0.25/0.75 as dashed lines, and 0.05/0.95 as dotted lines. *Column IV:* Confusion matrix for binary classification distinguishing the target condition from all other conditions at the same hierarchical level. **c,** (left to right) Demographic information for the exemplar subject; horizontal bar plots of z-scored morphological features representing the subject morphological profile; predicted label probabilities derived from the morphological profile, age, and sex (dashed lines indicate chance probability); the subject morphological profile overlaid with the top predicted diagnosis; the morphological feature space projected via UMAP for the top predicted diagnosis; and the second ranked alternative diagnosis, with the subject position marked. Note only relevant conditions are projected in UMAP space at each level in order to improve spatial resolution between relevant conditions.

**Extended Data Fig.2:**
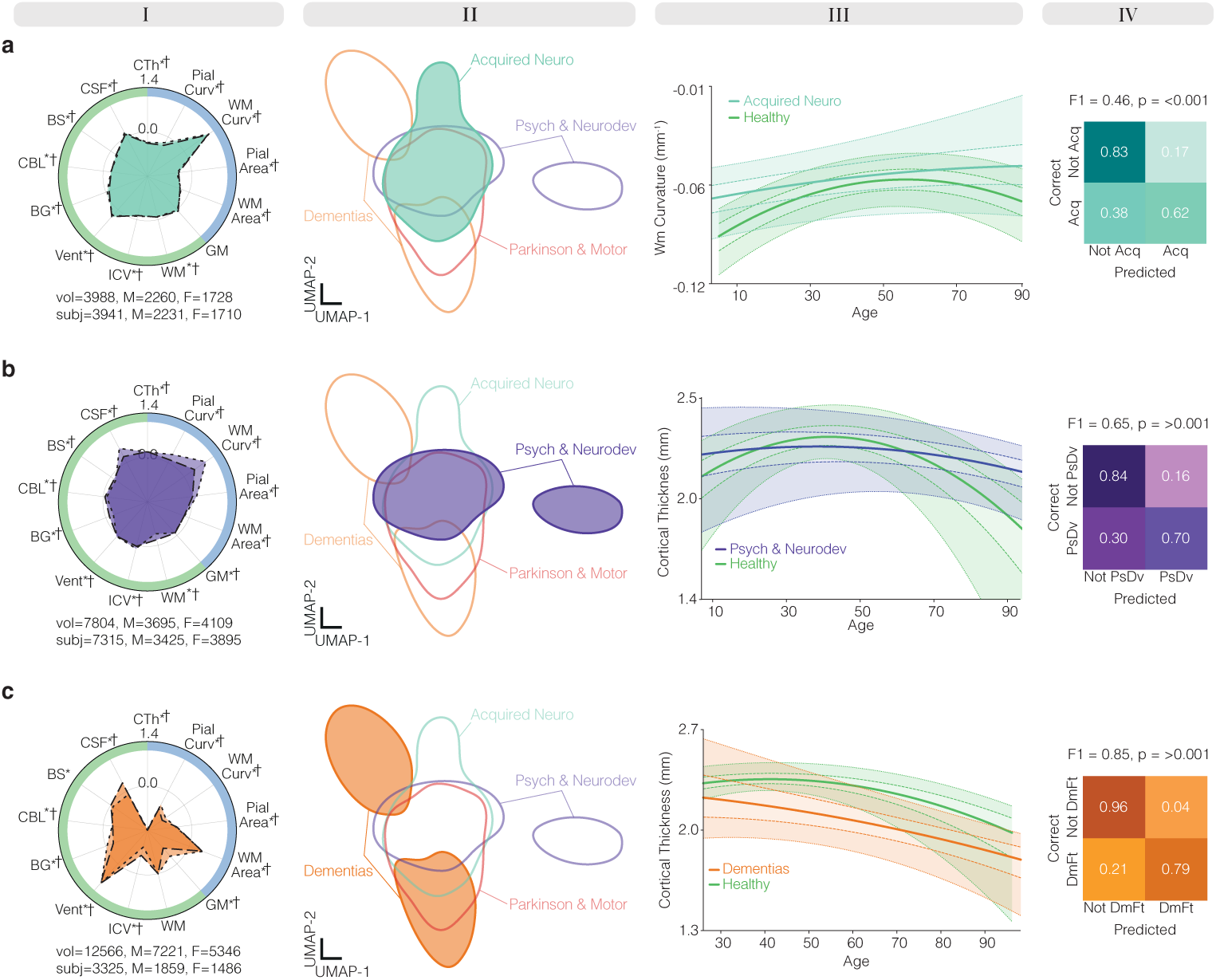
Morphological differentiation across families. Differentiation for other disease families including Acquired Neurological (**a**), Psychiatric and Neurodevelopmental (**b**) and Dementias families (**c**). *Column I:* Morphological feature profile of the target condition at the family level, represented as a radar plot. The centre circle represents the age and sex controlled healthy cohort. Significant differences between male and female cohorts are denoted by * and *†*, respectively. *Column II:* UMAP projection of the shared morphological feature space across conditions at the family level. *Column III:* Normative trajectories for the target condition relative to the healthy group across the age range of the target cohort. Normative centiles are represented as follows: 0.5 (median) as a bold line, 0.25/0.75 as dashed lines, and 0.05/0.95 as dotted lines. *Column IV:* Confusion matrix for binary classification distinguishing the target condition from all other conditions at the same hierarchical level.

We first summarized each group’s deviation from age- and sex-matched controls using radar plots (Fig. 3, col. I; Method 5). At the family level, deviations spanned almost all morphological descriptors, most prominently cerebralspinal fluid (CSF), ventricular volume and cerebellar volume, consistent with widespread atrophy. At the diagnosis level, PD showed a milder but still distributed morphological deviation profile. Within the morphology reference space, the family overlapped substantially with neighbouring families (Fig. 3, col. II; Method 3), and this overlap became more pronounced for PD, which sat among the other diagnoses within the same family. We then examined the age trajectory of the descriptor that best separated each group from the remainder of that level (Fig. 3, col. III; Method 2). White matter surface area distinguished the Parkinsonism and Motor family, whereas ventricular volume distinguished PD, and both deviated from the healthy trajectory across the age range examined. Although neighboring conditions partially overlapped within the reference space, quantitative classification confirmed that they remained distinguishable (col. IV, Method 4a). Using only the complete morphological profile set (thirteen feature profiles), the family was identified with a specificity of 79% and a sensitivity of 64%, and PD followed the same pattern, with a specificity of 87% and a sensitivity of 58%.

We finally examined whether an individual morphological deviation profile could recover this organization at the single-patient level (Fig. 3c). For a 63-year-old man with a clinical diagnosis of PD (col. I), his morphological deviation profile (col. II), together with age and sex, predicted the correct label at every level of the hierarchy: disease at the population level, Parkinsonism and Motor at the family level, and PD at the diagnosis level. CBS and PSP were returned as the next most probable alternatives (col. II), consistent with their clinical and morphological proximity within the reference space. A radar plot confirmed the alignment of his profile with the predicted PD signature (col. III), and in the shared space he fell within the PD cluster and outside that of CBS, his closest alternative (col IV). Additional profile reports for seven randomly sampled individuals are shown in Supplementary Fig 8.

Together, these results demonstrate that the morphology reference space supports differential characterization across clinically overlapping disorders by contextualizing individual patients relative to neighboring conditions. Although discrimination favored exclusion over confirmation, the resulting representation remained sufficiently informative to recover clinically meaningful assignments at the individual-patient level while identifying plausible alternative diagnoses.

### The Morphology Reference Space Captures Diagnostic Comorbidity

Diagnostic comorbidity poses a complementary challenge to differential characterization. Rather than asking which diagnosis best explains an individual’s morphology, it asks whether the same morphological representation contains evidence for more than one coexisting disorder. We therefore asked whether the morphology reference space could simultaneously represent multiple conditions within a single individual morphological deviation profile. Here, we focus on the Psychiatric and Neurodevelopmental family as a representative case, since autism spectrum disorder (ASD) and attention deficit hyperactivity disorder (ADHD) frequently co-occur [18]. Using the same analytical framework as for differential characterization, we examined morphology at the family and diagnosis levels (Fig. 4a,b), before turning to individual patients (Fig. 4c).

**Fig. 4:**
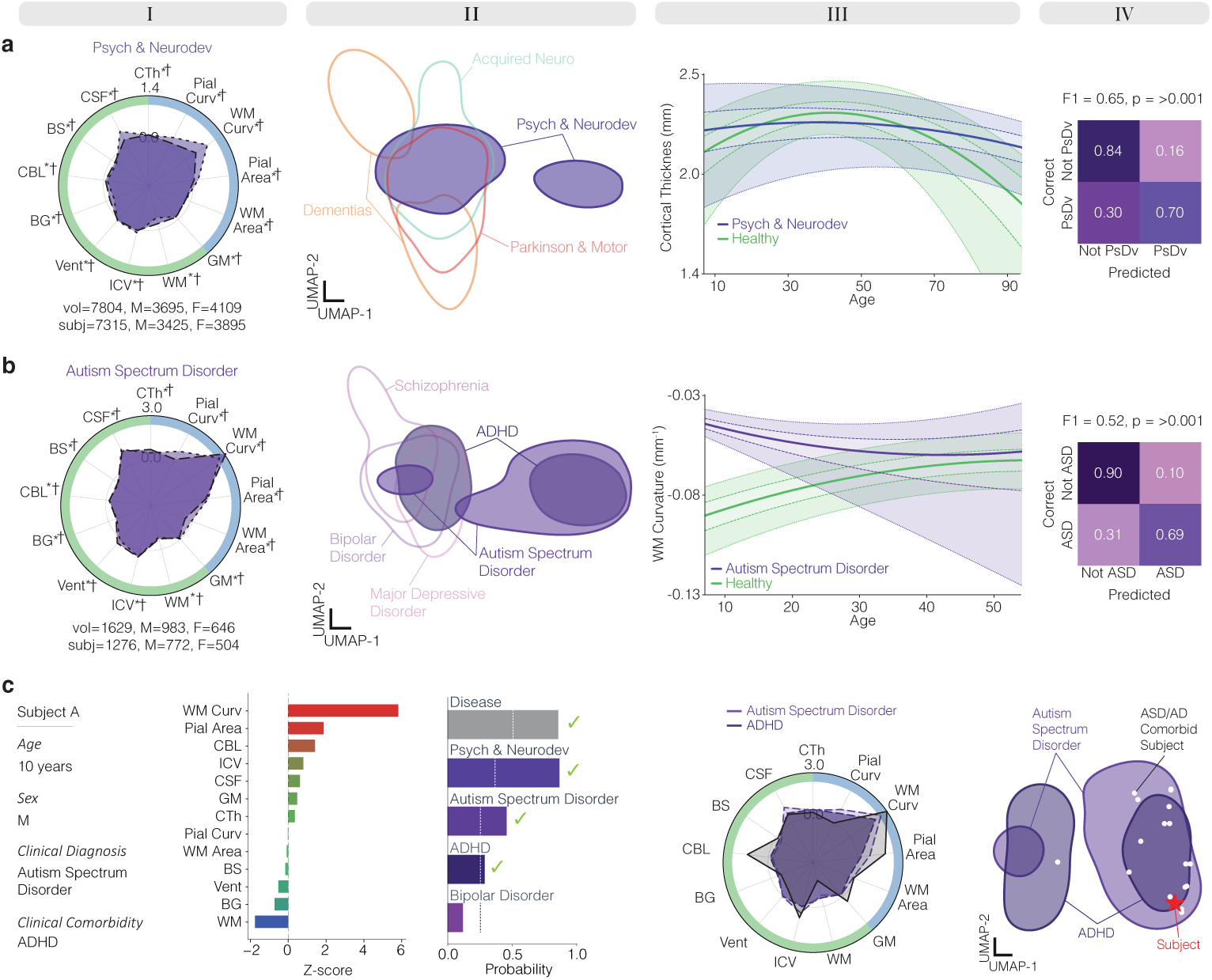
Morphological deviation profiles map diagnostic comorbidity. **a-b,** *Column I:* Group-level morphological deviation profile of the target condition at the family (**a**) and diagnosis (**b**) level, represented as a radar plot. The centre represents the age- and sex-matched healthy reference. Significant differences between male and female cohorts are denoted by * and *†*, respectively. *Column II:* UMAP visualization of the shared morphological deviation space across conditions at the family (**a**) and diagnosis (**b**) level. *Column III:* Normative trajectories of morphological descriptors for the target condition relative to the healthy group across the target cohort’s age range. Healthy normative centiles are represented as follows: 0.5 (median) as a bold line, 0.25/0.75 as dashed lines, and 0.05/0.95 as dotted lines. *Column IV:* Confusion matrix for binary classification distinguishing the target condition from all other conditions at the same hierarchical level. **c,** From left to right: demographic and clinical information for the exemplar subject; horizontal bar plots representing the subject morphological deviation profile; predicted label probabilities derived from the morphological profile, age, and sex (dashed lines indicate chance probability); subject profile overlaid with the top predicted diagnosis; and UMAP projection of the ADHD and ASD diagnostic space, with the subject position marked as a red star and ADHD/ASD comorbid participants marked as white dots. Only relevant conditions are projected into UMAP space at each level to improve spatial resolution among relevant conditions.

At the family level, morphological deviation profiles remained close to the healthy norm but showed distributed departures across curvature, cortical thickness, CSF and tissue volumes (Fig. 4a, col. I; Method 5). At the diagnosis level, ASD showed a stronger profile, dominated by white matter curvature, that partially overlapped that of ADHD (Fig. 4b, col. I). Within the morphology reference space, the Psychiatric and Neurodevelopmental family separated into two regions, a structure that persisted at the diagnosis level, where ASD and ADHD occupied neighboring, partially overlapping territories rather than isolated clusters (Fig. 4, col. II; Method 3). Age trajectories and classification confirmed that these profiles retained diagnostic signal despite this overlap, with both the family and ASD separated from the remainder of the hierarchy, again with higher specificity than sensitivity (Fig. 4, col. III-IV; Method 2).

We finally asked whether an individual morphological deviation profile could simultaneously support more than one diagnosis (Fig. 4c). For a 10-year-old boy with clinically confirmed ASD and ADHD, the profile predicted the disease group, the Psychiatric and Neurodevelopmental family, and ASD, while also returning ADHD as a secondary above-threshold prediction, consistent with the patient’s clinically confirmed comorbidity. His radar profile aligned with both ASD and ADHD signatures, and his position in the shared space fell within their overlap region. Importantly, this observation generalized across the clinically confirmed comorbid cohort, whose patients consistently occupied the ASD-ADHD overlap region rather than scattering between the two conditions.

Together, these results demonstrate that the morphology reference space naturally captures diagnostic comorbidity by positioning patients within regions shared by multiple disorders, extending beyond single-label classification toward anatomically grounded clinical interpretation.

### Longitudinal Trajectories Within The Morphology Reference Space

The previous analyses characterized the morphology reference space at a single point in time. A further property of this space is that individuals can be followed longitudinally, transforming a static representation into a dynamic trajectory of disease progression. We therefore focused on the Dementias family, where progression from healthy ageing to mild cognitive impairment (MCI) and Alzheimer’s disease (AD) is routinely studied [19].

At the family level, deviations from controls were pronounced across all features, reflecting widespread atrophy and cortical change (Fig. 5a, col. I; Method 5). Within the morphology reference space, the family split into two distinct regions, with some individuals blending into neighboring groups and others forming an isolated cluster (Fig. 5a, col. II; Method 3). The trajectory of the most discriminative descriptor for each group diverged sharply from the healthy trajectory across the age range (Fig. 5a, col. III; Method 2), contrasting with the weaker departure observed in the Parkinsonism and Motor family. This structure yielded the most accurate family classification we observed, ruling the family out in 96% of cases and confirming it in 79% (Fig. 5a, col. IV; Method 4a). At the diagnosis level, AD and MCI overlapped considerably, as did FTD and PPA, while the two pairs remained distinct; AD was again ruled out more readily than confirmed, at 78% versus 67% (Fig. 5b).

**Fig. 5:**
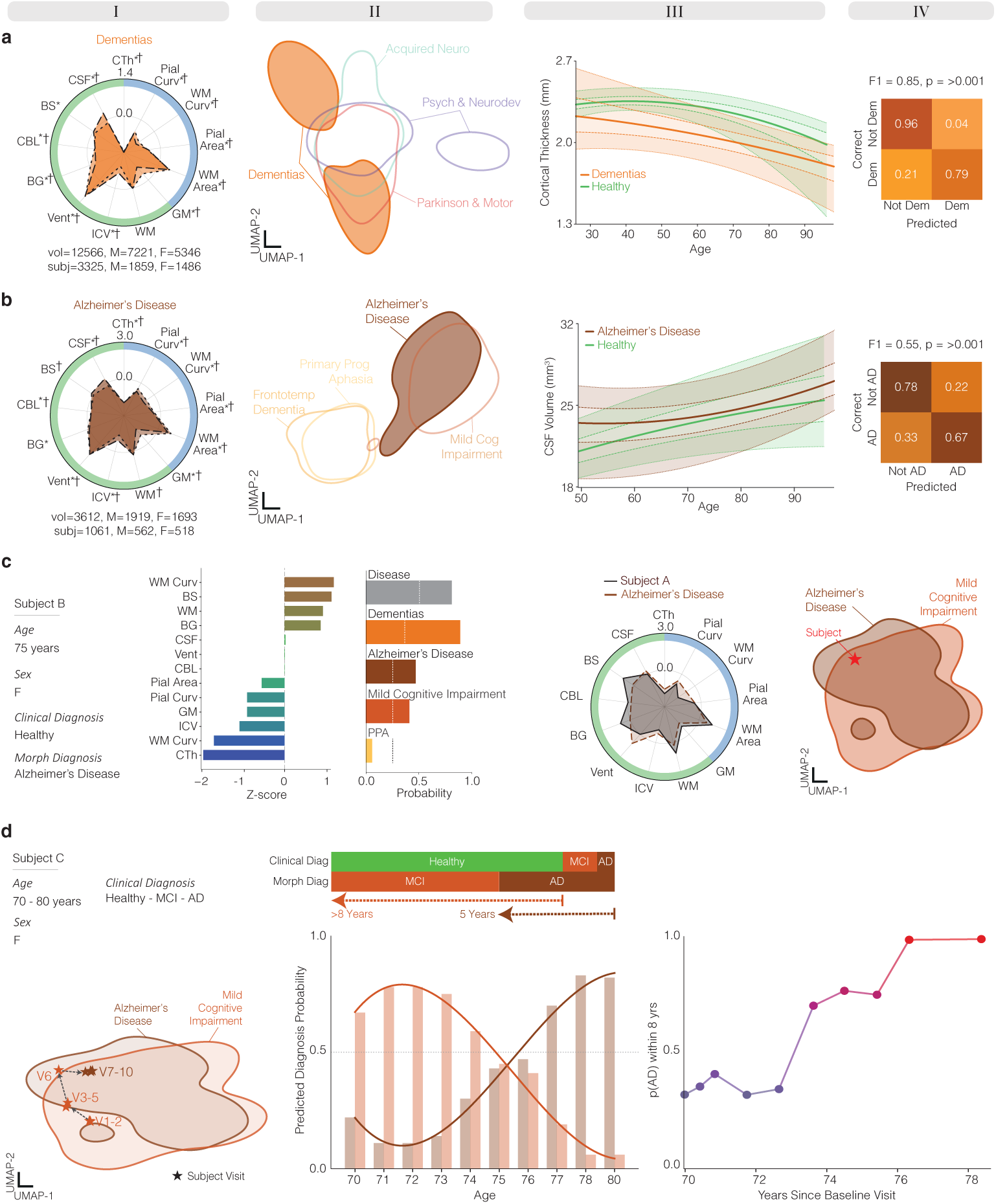
Longitudinal morphological trajectories. **a-b,** Differential morphological profiling of the Dementias family (**a**) and Alzheimer’s Disease diagnosis (**b**) including radar profile (*Column I*), shared morphological space (*Column II*), normative modeling of the most predictive feature (*Column III*) and binary classification confusion matrix (col IV). **c** Individual morphological profiling of a 75-year-old female with a healthy clinical diagnosis but an AD morphological profile. Individual profile includes (from left to right) subject demographics, morphological feature deviations, label prediction, individual profile compared to top predicted diagnosis profile, and shared morphological space projected in UMAP space for top two diagnoses. **d** Longitudinal morphological profiling of the same individual across ten annual visits, showing her position in the shared morphological space at each visit, the corresponding predicted label probabilities over time, and t^1^h^5^e Cox-modeled probability of developing AD within an eight-year window.

We next asked whether this organization could also describe disease progression within an individual patient (Fig. 5c; Method 6). In a woman in her mid-seventies, baseline clinical assessment indicated healthy status, yet her morphology already showed disease-related deviation, placing her within the Dementias family with similar probabilities assigned to AD and MCI. (Fig. 5c). Across ten consecutive years of imaging, her position progressively traversed the morphology reference space, moving from the MCI region, through its overlap with AD, and into the AD region, while diagnostic probabilities shifted in parallel (Fig. 5d). This anatomical progression was mirrored by Cox proportional hazards models fitted at each visit (Method 7), with the estimated eight-year AD risk increasing alongside her movement through the reference space.

This case illustrates how a morphology reference space constructed from population data can support individual patient interpretation over time by locating, ordering and monitoring progression through the landscape of brain disease.

## Discussion

We established a shared morphology reference space that positions individual brains within the broader organization of neurological and psychiatric disease, providing a common anatomical context for interpreting structural MRI across disorders. The space was constructed from 110,591 sMRI scans spanning four disease families, 19 diagnoses and seven subtypes. It moves beyond conventional case-control morphometry, which asks whether disease groups differ from healthy controls, and extends normative modeling, which asks whether an individual differs from age- and sex-appropriate expectations, by asking what that individual’s morphology resembles across the broader landscape of brain disease. Its principal advance is therefore not the provision of another disease classifier, but the construction of a reference coordinate system in which morphological relationships among healthy populations, disease families, diagnoses, subtypes and individual brains can be examined directly. This distinction is substantive rather than semantic. Whereas a conventional classifier returns a single categorical label, the morphology reference space places that label within a broader relational context. Positioning an individual within the hierarchical space identifies their morphological neighbors, quantifies overlap with adjacent or comorbid conditions and, when assessed longitudinally, traces their trajectory of morphological change. This position can be interpreted through compact profiles of individual morphological deviation, with diagnostic assignments accompanied by corresponding estimates of uncertainty. Together, these findings advance individualized morphometry from identifying whether an individual brain is atypical, to determining what that atypicality resembles across the wider landscape of brain disease, providing a scalable foundation for relational neuroimaging, cross-disease phenotyping and patient-level profiling.

Structural neuroimaging has characterized disease-associated morphology in considerable detail, but most studies have examined individual disorders in isolation, either through group-level case-control comparisons or normative models developed for selected diagnoses and clinical endpoints [7, 8]. Large-scale brain charts and normative modeling have provided essential references for assessing whether an individual brain deviates from expected variation [4, 6], while transdiagnostic and classification studies have compared selected conditions within predefined analytical tasks [11]. The morphology reference space connects these approaches by deriving every individual morphological deviation profile from the same healthy reference and positioning it within a common hierarchy spanning disease families, diagnoses and subtypes. Consequently, morphological resemblance between conditions becomes a directly measurable property of the space rather than an inference assembled across separately acquired cohorts, analytical pipelines and published contrasts. The framework therefore shifts the emphasis from identifying isolated disease effects or optimizing a single diagnostic distinction, towards mapping how individual deviations relate across the broader landscape of brain conditions.

Across the hierarchy, morphology retained discriminative structure from population and disease family to diagnosis and subtype, although overlap increased as the distinctions became progressively finer. This pattern suggests that broad disease families exhibit relatively coherent forms of neuroanatomical involvement, whereas diagnostic and subtype boundaries are less anatomically discrete. Importantly, this overlap should not necessarily be interpreted as noise or model failure. Rather, it may reflect shared structural consequences, heterogeneity within clinical diagnoses, differences in disease stage or distinct pathological processes converging on similar macroscopic anatomy. The comparatively strong separation observed at the disease-family level may therefore indicate that brain morphology captures broad patterns of neuroanatomical involvement more faithfully than every boundary imposed by categorical diagnosis. Notably, the hierarchy was clinically defined rather than discovered from morphology de novo; these findings therefore show that brain morphology recapitulates clinically meaningful organization, not that it establishes a new biological taxonomy of disease.

Throughout the hierarchy, the strongest discrimination arose from the combined contribution of multiple morphological descriptors rather than from any single measurement. This indicates that disease morphology is better represented through the relative configuration of complementary anatomical properties, rather than through any single marker such as cortical thickness, tissue volume or brain age gap [20]. NeuroMorph captures this configuration as a compact morphological fingerprint, while normative modeling transforms it into an interpretable morphological deviation profile by quantifying how each component departs from age- and sex-appropriate expectations. The resulting 13-descriptor representation occupies a deliberate middle ground. Rather than using atlas-based cortical parcellations comprising hundreds or thousands of correlated regional measurements, we prioritized a compact set of complementary whole-brain descriptors that could be compared consistently across conditions, interpreted at the individual level and applied at population scale. This design retains more information than any individual descriptor, while reducing dependence on atlas selection and preserving interpretability across heterogeneous datasets. The representation therefore prioritizes robust positioning within the morphology reference space over fine-grained anatomical localization. Regional extensions may provide additional spatial specificity for applications in which localization itself is the primary objective, while preserving the same conceptual framework.

The three individual-level applications illustrate complementary ways in which the same morphology reference space can be interrogated to contextualize clinical complexity. In the differential characterization example, the morphological deviation profile of a patient with Parkinson’s disease aligned most closely with the clinically assigned diagnosis while retaining similarity to progressive supranuclear palsy and corticobasal syndrome. This example illustrates how morphology could help organize a set of plausible diagnostic alternatives rather than force an unequivocal label. Comorbidity represents a complementary form of diagnostic ambiguity: individuals with co-occurring ASD and ADHD occupied regions associated with both conditions, indicating that an individual morphological profile can retain resemblance to more than one diagnostic group rather than being reduced to a single category. This should not be interpreted as evidence of shared etiology, but as a measurable convergence in their macroscopic anatomical expression. Longitudinally, repeated positioning transformed the morphology reference space from a static representation into an individual disease trajectory, with one participant moving from healthy status towards MCI and Alzheimer’s disease as diagnostic probabilities and estimated risk increased in parallel. Although these examples are illustrative and require cohort-level validation, together they demonstrate that the same morphology reference space can support three complementary forms of patient interpretation that categorical diagnosis captures only incompletely: similarity between competing diagnoses, coexistence of overlapping conditions, and disease progression over time.

The consistent asymmetry between exclusion and confirmation further suggests that the near-term role of the morphological reference space may be to rule out morphologically inconsistent alternatives or flag cases requiring more specific clinical, molecular or imaging assessment, rather than to replace diagnostic judgment. Its use of routine T1-weighted MRI, rapid processing and compact individual-level outputs could support integration into large-scale clinical imaging pathways, longitudinal follow-up and trial stratification. Clinical deployment, however, will require prospective validation, calibrated probabilities, comparison with existing clinical assessment and demonstration that morphological profiling adds actionable information beyond standard care.

The practical utility of a morphology reference space depends on its ability to be applied consistently across datasets, imaging environments and individual patients. NeuroMorph derives each morphological fingerprint from a routine T1-weighted MRI scan within seconds and operates in native anatomical space, preserving individual geometry without requiring computationally intensive surface-processing pipelines. This enables large-scale construction and repeated application of the reference space using routine clinical imaging. Because morphological deviation profiles are expressed relative to age- and sex-appropriate normative expectations, individual subjects can be interpreted without assembling a new matched control group at the point of analysis. The framework also showed robustness across scanners, sites and heterogeneous acquisition protocols, enabling the same representation to be applied consistently from population-scale cohorts to repeated measurements within a single patient. Together, rapid processing, routine clinical inputs and standardized individual-level outputs make the construction and deployment of a large cross-disease reference space feasible in both research and clinical settings.

Several limitations should be considered when interpreting these findings. The individual-level findings are illustrative rather than confirmatory, and the comorbidity result is drawn from a single family of conditions, providing an initial demonstration rather than a general conclusion across neurological and psychiatric disorders. Diagnostic labels throughout are clinically defined rather than pathologically confirmed, and for several of the conditions we sought to separate, clinical diagnosis is itself an imperfect reference standard [16]. Differences in the granularity and consistency of contributing cohorts constrain the resolution at which conditions can be separated, most notably for the subtypes with the smallest samples. The UMAP projections used for visualization summarize a much higher-dimensional space into two dimensions and were not used for quantitative inference. They should therefore be interpreted as qualitative visualizations of the underlying organization rather than as quantitative measures of biological proximity or separability.

Future work should determine whether the disease morphology reference space captures diagnostic comorbidity more generally across neurological and psychiatric disorders, establishing whether localized overlap represents a common property of disease morphology or whether the ASD-ADHD pattern is specific to neurodevelopmental conditions. The same reference space may also reveal morphological subtypes that cut across conventional diagnostic boundaries and support disease staging within individual conditions [21]. Validation against pathological, molecular or fluid biomarkers will be important to determine whether the observed organization reflects shared disease biology or convergent structural consequences. Integrating morphology with PET, blood-based biomarkers, CSF, genetics and cognitive measures may further enrich the reference space, enabling multimodal representations with greater diagnostic and prognostic specificity. Ultimately, prospective external validation will be required to establish whether longitudinal movement within the morphology reference space improves patient stratification, predicts clinically meaningful outcomes and supports treatment selection or clinical trial enrichment. Finally, future work should investigate the influence of other modifiable factors known to affect brain structure, including socioeconomic status and lifestyle [55].

## Methods

### Data and Preprocessing

We analysed 25 datasets comprising 110,591 MRI volumes (see Supplementary section *’Datasets’* for a comprehensive list of datasets, including number of volumes and participants). The inclusion of data from 25 independent cohorts introduced substantial variability in acquisition protocols and scanners, reflecting real-world clinical heterogeneity. All T1-weighted images were conformed to 256^3^ at 1 mm isotropic resolution in LIA orientation and intensity-standardized via within-volume z-scoring. No additional preprocessing, such as skull stripping, bias-field correction or registration to MNI space was applied. Operating on native-space images minimized preprocessing-induced changes to morphology and intensity (including interpolation artefacts), preserved subject-specific anatomical variability and enabled consistent estimation of morphology across heterogeneous datasets, thereby supporting construction of the morphology reference space at population scale. For all analyses, we included only T1-weighted MRI volumes from participants aged 3–100 years. All scans underwent dataset-level quality control as defined in the original studies. In addition, we screened the derived 13-feature morphological fingerprint for anomalous values. Volumes in which multiple features were identified as extreme outliers relative to the remainder of the dataset were flagged for manual review and excluded if the image was judged to be corrupt or incomplete.

Data span 19 neurological and psychiatric disease diagnoses, including Alzheimer’s Disease (AD), Mild Cognitive Impairment (MCI), Frontotemporal Dementia (FTD), Primary Progressive Aphasia (PPA), Parkinson’s Disease (PD), Progressive Supranuclear Palsy (PSP), Corticobasal Syndrome (CBS), Motor Neuron Disease (MND), Multiple Sclerosis (MS), Meningitis (MNG), Schizophrenia (SCHZ), Bipolar Disorder (BP), Major Depressive Disorder (MDD), Autism Spectrum Disorder (ASD), Attention Deficit Hyperactivity Disorder (ADHD), Traumatic Brain Injury (TBI), Epilepsy (EPI), Stroke, and Tumour. Together with healthy controls (HC), this yielded 20 diagnostic categories in the aggregated cohort (Fig. 6 a,b). Stroke and Tumour labels reflect clinically defined events and do not distinguish subtypes (for example, haemorrhagic vs ischaemic stroke, or tumour histology). Diagnostic labels were taken directly from the source datasets and followed the clinical criteria defined by each contributing study. Volumes or participants without a diagnostic label were excluded from predictive analyses, but were retained for brain mapping analyses when age and sex were available. Sex was approximately balanced across the overall cohort; however, the age distribution was skewed towards older individuals, with most participants between 50 and 80 years, consistent with the predominance of late-onset neurodegenerative disorders in the cohort (Fig. 6c,d).

**Fig. 6:**
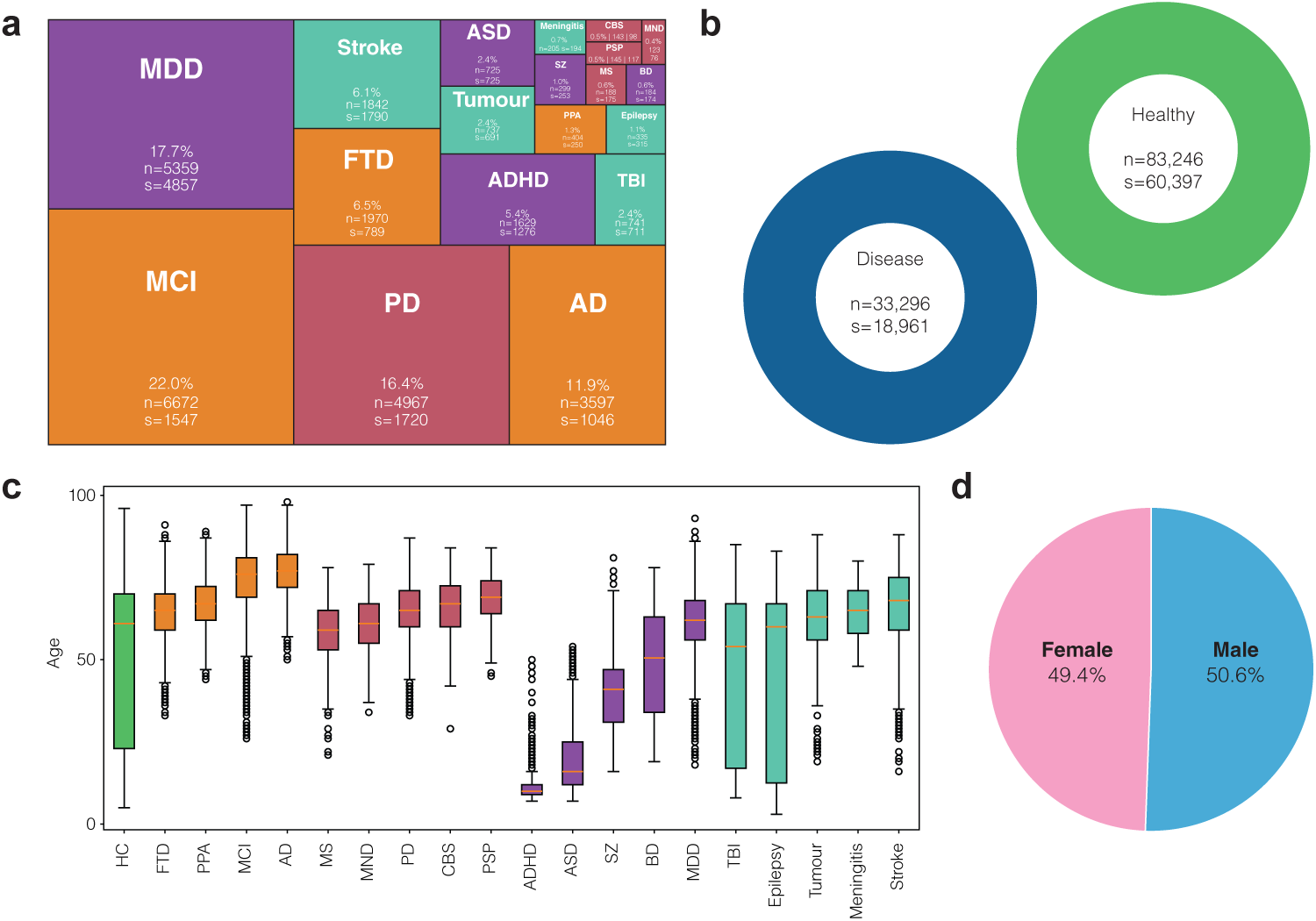
Data Distributions. **a,** Treemap showing each diagnostic group’s contribution to the overall disease cohort; tiles are annotated with percentage contribution, number of volumes (n) and number of participants (s). **b,** Donut charts summarising the disease and unhealthy cohorts, annotated with the number of volumes (n) and participants (s). **c,** Box plots showing age (years) by diagnostic group; colors indicate the diagnostic family for each group. **d,** Sex distribution across diagnostic groups.

### Datasets

Datasets include Adolescent Brain Cognitive Development (ABCD) Study (n vol = 19059, participants = 9659) [22], AIMSTBI (n vol = 388, participants = 388) [23], ARTFL LEFFTDS Longitudinal Frontotemporal Lobar Degeneration (ALLFTD) (n vol = 3784, participants = 1833) [24], the Amsterdam Open MRI Collection(AOMIC) (n vol = 1760, participants = 865) [25], BrainLat (n vol = 572, participants = 571) [26], ABIDE (Autism Brain Imaging Data Exchange) ((n vol = 975, participants = 975)) [27], ADHD200 (n vol = 772, participants = 772) [28], GLASGOW (n vol = 1199, participants = 1199), HCP (n vol = 874, participants = 874) [29], IDEAS (n vol = 277, participants = 277) [30] , IXI (n vol = 536, participants = 536) (see acknowledgements), Mayo Clinic Study of Aging (MCSA) (n vol = 6180, participants = 3089) [31], Frontotemporal Lobar Degeneration Neuroimaging Initiative (NIFD) (n vol = 1621, participants = 300) (http://memory.ucsf.edu/research/studies/nifd, see acknowledgements), OASIS3 (n vol = 2537, participants = 1194) [32], Parkinson’s Progression Markers Initiative (PPMI) (n vol = 5386, participants = 1759) (see acknowledgements), Stroke Outcome Optimization Project (SOOP) (n vol = 1106, participants = 1106) [33], SRPBS (n vol = 1383, participants = 1383) [34], UCLA Neuropsych (n vol = 265, participants = 265) [35], The UK Biobank (UKB) (n vol = 53645, participants= 44451) [36], The University of Pennsylvania glioblastoma (UPenn-GBM) cohort (n vol = 671, participants = 630) [37], apetMND (n vol = 118, participants = 59) [38], eFCD2 (n vol = 170, participants = 170) [39], solervidal (n vol = 142, participants = 71) [40] and the Alzheimer’s Disease Neuroimaging Initiative (ADNI) (n vol = 13122, participants = 2011) (adni.loni.usc.edu, see acknowledgements). Data used in the preparation of this article were obtained from the Alzheimer’s Disease Neuroimaging Initiative (ADNI) database (adni.loni.usc.edu). The ADNI was launched in 2003 as a public-private partnership, led by Principal Investigator Michael W. Weiner, MD. The primary goal of ADNI has been to test whether serial magnetic resonance imaging (MRI), positron emission tomography (PET), other biological markers, and clinical and neuropsychological assessment can be combined to measure the progression of mild cognitive impairment (MCI) and early Alzheimer’s disease (AD). A breakdown of how each dataset contributes to the healthy group and each diagnosis group can be observed in Supplementary Fig. 9 and Supplementary Fig. 10, respectively.

### Method 1: NeuroMorph - A Deep Learning Framework for Morphological Fingerprinting

NeuroMorph (Supplementary Fig. 11) combines LOD-Brain^+^ (Method 1a) and DeepThickness (Method 1b) to derive eight volumetric and five surface-based features, yielding a 13-feature morphological fingerprint from a single T1-weighted MRI scan. Unless stated otherwise, all references to morphological features and the morphological fingerprint in the Results refer to outputs generated by this framework. All T1w MR images were conformed to a 256^3^ voxel grid in LIA orientation and intensity-normalized using z-score standardization. No additional preprocessing was performed. Images were not registered or normalized to MNI space, and no bias field correction was applied.

### Method 1a: Volumetric Feature Extraction

Eight volumetric features were derived using an adapted version of LOD-Brain [41], hereafter referred to as LOD-Brain^+^, which generates an eight-label tissue segmentation (gray matter, white matter, intracranial, ventricular, cerebellar, brainstem, basal ganglia and cerebrospinal fluid volumes) from T1-weighted MRI volumes. The original LOD-Brain model, builds on previous work [42, 43], using a fully 3D convolutional neural network designed for robust MRI segmentation across heterogeneous scanners and acquisition sites. It employs a progressive level-of-detail architecture in which U-Net based subnetworks operate at increasing spatial resolutions, segmenting from coarse to fine scale. The original model outputs seven anatomical classes, operates directly on minimally pre-processed T1w MRI volumes without atlas registration, fine-tuning, or site-specific adaptation, and was trained on a large multi-site dataset using weak supervision derived from FreeSurfer segmentations [41]. In the present study, we retrained the original LOD-Brain model to include an explicit CSF mask, yielding an eight-class segmentation. Because FreeSurfer does not provide a dedicated CSF label, we generated the CSF reference mask with SynthSeg [44] and used FreeSurfer-derived labels for the remaining seven reference classes. These combined labels were used as supervision to retrain the model to predict all eight classes. Retraining used 1,049 training volumes and 75 validation volumes from 11 datasets (Supplementary Fig. 12). The retrained model was subsequently applied to all MRI volumes, from which quantitative volumetric descriptors were computed. The volumes used to retrain LOD-Brain were drawn from the same aggregated cohort used in the main analysis. As these volumes contributed only to the tissue segmentation task rather than the downstream diagnostic or predictive analyses, their reuse does not constitute data leakage relevant to the main findings. For all analyses (except trajectory-based visualization), volumetric features were normalized by total intracranial volume (ICV). This approach is supported by evidence that volumetric normalization to ICV can enhance sensitivity to relatively subtle neurodegenerative effects that may otherwise be obscured by variance attributable to global cranial size [45]. For a more detailed description of the original model, including training evaluation, robustness to site and scanner effects, and comparison to other methods such as FreeSurfer, see [41].

### Method 1b: Surface Feature Extraction

#### Overview

Five surface-based features (cortical thickness, pial curvature, white matter (WM) curvature, pial surface area, and WM surface area) were derived using *DeepThickness*, a deep learning-based 3D U-Net developed for cortical thickness estimation and surface reconstruction. DeepThickness takes as input an isotropic native-space T1w volume together with the corresponding tissue segmentation and white- and gray-matter probability maps generated with LOD-Brain^+^. The network simultaneously predicts a level-set volume defining the target surface and a cortical thickness (CTh) distance-field volume, conditioned on the requested surface type. Reconstructed meshes were post-processed and projected onto the CTh distance field to generate vertex-wise CTh overlays for the WM and pial surfaces.

#### Data, preprocessing, and augmentation

The model was trained on 1,049 T1w MRI volumes from mixed clinical and research cohorts across multiple scanners (Supplementary Fig. 13), with performance monitored on a validation set of 130 cases comprising 53% internal and 47% external data. Data augmentation was applied exclusively to the training set. To improve robustness and generalization, stochastic augmentation was applied to T1w MRI volumes during training, following the procedure outlined in [41].

#### Model

The DeepThickness network (Supplementary Fig. 14) follows a 3D U-Net style encoder-decoder architecture [46] designed to learn cortical surface representations from T1-weighted MRI input, with skip connections preserving fine anatomical detail across resolution levels. Feature extraction uses residual bottleneck blocks with 3 × 3 × 3 kernels, normalization, activation, and dropout, with downsampling and upsampling performed via strided convolutions rather than pooling. The decoder shares a common pathway before splitting into two task-specific branches that independently predict a level-set representation and distance set for the WM and pial surfaces. This *shared then split* design promotes consistency between surface parameterizations and improves parameter efficiency relative to training separate single-output networks.

#### Level Set Loss

Training used a custom ranged weighted mean absolute error loss combined with a Kullback-Leibler divergence term for distributional regularization. Voxel-wise weights emphasized a target range corresponding to a narrow band around the level set from which the surface boundary is extracted, with in-range voxels assigned an elevated weight of 10.0 and remaining voxels weighted according to their distance from zero, subject to a minimum of 0.01 to prevent vanishing gradients. The weighted MAE term was normalized to keep loss magnitude comparable across batches with differing proportions of in-range voxels, and the KL term compared normalized histograms of predicted and true values over the focus range. The total loss was defined as

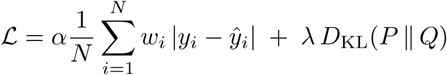

#### Distance Set Loss

Training used a capped weighted MAE loss to address class imbalance arising from a high proportion of uninformative or saturated voxels. Voxels whose ground truth values matched a predefined capped value, such as zero-valued voxels in distance maps, were assigned a reduced weight, while all other voxels received a higher weight to emphasize informative regions. The loss was normalized to remain comparable across batches with varying proportions of capped voxels. Capped voxels were weighted at zero in the reported experiments, effectively excluding them from the loss. The total loss was defined as

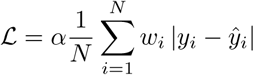

#### Model Training

Optimization used Adam [47] for a maximum of 50 epochs, starting from a learning rate of 0.0013 that was reduced by a factor of 0.25 upon validation plateau, with ELU activations, group normalization, and dropout at 0.1 applied throughout the encoder and decoder. Training ran on an NVIDIA A100 SXM4 GPU (80 GB), taking approximately 14 h 40 min for the pial surface and 11 h 42 min for the WM surface. Optimal configuration was determined through an ablation study (Supplementary Fig. 15) evaluating architectural and training choices, including the number of hierarchical levels, plain versus residual convolutional blocks, normalization strategy, and the balance between downsampling and upsampling versus repeating convolutional blocks across identity layers. Model selection was based on statistical significance on the validation set, with preference given to fewer trainable parameters when differences were not significant.

#### Model Evaluation

Model performance was assessed using metrics evaluating the predicted level and distance sets (model loss, mean squared error (MSE), and mean absolute error (MAE)) and the resulting mesh (ASSD, Hausdorff, Hausdorff 90, and number of intersections) for both pial and WM representations. As shown in Supplementary Fig. 16, performance was comparable between validation samples drawn from the same source datasets used during training (internal validation) and samples originating from independent datasets not included in training (external validation), indicating robust generalization across heterogeneous data sources (e.g. site and scanner effects).

#### Cortical surface reconstruction

The level-set representation encodes a surface implicitly as the zero level set of a scalar field, from which a triangulated mesh is recovered using the marching cubes algorithm [48], followed by postprocessing to remove erroneous bridges between gyri and achieve the desired sulcal depth. Surfaces were decimated to 7,500 vertices and smoothed using Taubin and Laplacian methods [49–51] to reduce common reconstruction irregularities. Cortical thickness was then represented as a distance set, a three-dimensional scalar field that is zero everywhere except at voxels intersected by the cortical surfaces, where values encode the local distance between the inner and outer boundaries. These values were computed by linearly interpolating surface vertices into the image volume using SciPy’s RegularGridInterpolator and assigning the corresponding interpolated distance values. Examples of cortical surface reconstructions with CTh overlay generated by DeepThickness compared to FreeSurfer are shown in the Supplementary Fig. 17.

### Method 1c: NeuroMorph Evaluation

To establish the suitability of NeuroMorph for large-scale and heterogeneous neuroimaging studies, we evaluated its agreement with the widely used FreeSurfer framework [52], together with its anatomical fidelity and computational efficiency. This evaluation comprised three complementary benchmarks: (a) quantitative agreement, (b) qualitative alignment, and (c) computational efficiency. We quantified agreement in derived volumetric and surface-based measures across both research and clinical datasets to assess measurement consistency. Next, we assessed anatomical plausibility by visually inspecting cortical segmentations and surface placement relative to the underlying T1-weighted MRI in representative participants. Notably, we conducted a blinded expert rating experiment in which neuroimaging specialists directly compared NeuroMorph and FreeSurfer surface reconstructions and selected their preferred output, providing an independent, clinically grounded assessment beyond automated quantitative metrics. Finally, we quantified computational efficiency by measuring inference time, enabling direct comparison of throughput relative to FreeSurfer.

**a) Quantitative agreement** Firstly, we evaluated quantitative agreement between NeuroMorph and FreeSurfer across all morphometric measures included in the fingerprint using the DeepThickness model validation set (130 T1-weighted MRI volumes drawn from 10 independent datasets; Supplementary Fig. 18). Across datasets, NeuroMorph showed strong agreement with FreeSurfer across most measures, with particularly high concordance for cortical thickness; the mean between methods difference was 0.07 mm, substantially smaller than the voxel resolution (1.0 mm). Notably, agreement was maintained in both clinical and research datasets (Supplementary Fig. 19). Despite this overall agreement, systematic shifts were observed. NeuroMorph yielded higher grey matter (GM) and correspondingly lower WM volume estimates, potentially consistent with a sharper delineation of the GM and WM boundary (Supplementary Figs 20, 21). The largest between-method differences were observed for mean curvature on both the WM and pial surfaces, which we attribute to differences in surface postprocessing and smoothing, and to localized FreeSurfer segmentation failures and topological artefacts that NeuroMorph reduced.
**b) Qualitative alignment** We assessed anatomical alignment by visually inspecting representative voxel-space examples (Supplementary Fig. 20). Across participants, NeuroMorph produced consistently more anatomically accurate cortical segmentations and closer surface alignment to the underlying MRI than FreeSurfer, and ameliorated several recurrent segmentation and surface-placement inaccuracies observed systematically across participants (Supplementary Fig. 21). This qualitative improvement was corroborated by a blinded expert neuroimaging evaluation of pial surface placement relative to the T1-weighted MRI, with NeuroMorph consistently preferred over FreeSurfer in both clinical and research datasets and across all cortical regions examined (Supplementary Fig. 22).
**c) Timing and Reliability** Finally, NeuroMorph provided a substantial computational advantage over FreeSurfer. Under the same execution conditions, with both pipelines run sequentially and CPU usage limited to four cores, mean processing time was reduced by 99.5%, from 3.5 hours per volume with FreeSurfer’s recon-all to 1.1 minutes per volume with NeuroMorph (Supplementary Fig. 23). Notably, FreeSurfer failed on 49 cases, whereas NeuroMorph succeeded on all volumes (Supplementary Fig. 24). In a separate timing experiment using NeuroMorph’s multi-threaded pipeline on 48 CPU cores (Intel Xeon Gold 6226, 2.70 GHz, two sockets of 24 physical cores each) and a single NVIDIA Quadro RTX 8000 GPU (48 GB VRAM), mean inference time fell further to 13 seconds per volume, averaged across 100 volumes. This additional gain in throughput further improves the practicality of large-scale neuroimaging analyses.

Overall, NeuroMorph produces morphometric measurements that are closely aligned with FreeSurfer-derived estimates while reducing segmentation and surface-reconstruction errors and substantially accelerating processing.

### Method 2: Normative Modeling for Morphological Features

To characterize deviations in brain morphology, we used normative modeling to construct a reference model of healthy cortical morphology and express each individual’s profile as a standardized deviation (z score) from this reference. Models were fitted using PCNtoolkit [53] (version 1.2.0). The healthy reference cohort comprised 79,674 of 110,591 total volumes, with the remaining 30,917 volumes belonging to individuals with a clinical diagnosis. The healthy cohort was partitioned at the subject level into a training set (95%) and a held out test set (5%), with age included as a covariate and sex modeled as a batch effect for each of the 13 morphological descriptors. Each descriptor was modeled independently using a Bayesian linear regression template in PCNtoolkit, with a cubic B spline basis (five knots) to capture nonlinear age effects, heteroskedastic noise to allow variance to vary with age, and a sinh arcsinh warp to account for departures from Gaussian distributions in the raw values. Inputs and outputs were standardized prior to fitting. During normative model fitting, 2,121 volumes exceeding an absolute z-score threshold of 5 were excluded to prevent extreme outliers from influencing model estimation. The fitted model was subsequently applied to all 108,470 volumes irrespective of diagnostic status, generating a z score per subject and morphological descriptor that reflected the deviation of the observed value from that expected given the subject’s age and sex. These subject-specific z-scores collectively define the morphological deviation profile used throughout the Results.

### Method 3: Visualization of the Morphology Reference Space

For visualization purposes, participants were projected into a two-dimensional space using Uniform Manifold Approximation and Projection (UMAP) [54], applied to their morphological deviation profiles. For each diagnostic grouping considered, the spatial extent of every group within the embedding was characterized using Gaussian kernel density estimation, with bandwidth set according to Scott’s rule and scaled by a fixed factor. Density was evaluated at the location of every subject belonging to the group, and the value at a chosen percentile of these densities was taken as a single contour threshold, producing a density contour that approximately encloses the participants associated with the highest local density for that group while excluding sparsely populated outlying regions. These contours provide a qualitative representation of the distribution of each group within the embedding and do not represent the output of a clustering algorithm or a quantitative measure of biological proximity or separability. Embedding outliers were excluded prior to boundary estimation using an interquartile range filter applied independently to each embedding dimension. Throughout the manuscript, UMAP embeddings are used exclusively for visualization. All quantitative analyses were performed in the original 13- dimensional morphology reference space. This approach was applied at three levels of diagnostic grouping, described below.

#### Binary clinical grouping

To visualize the separation between affected and unaffected individuals, participants were first downsampled to an equal number of 5000 per clinical class (random state 42), removing the influence of unequal class sizes on the resulting embedding and boundaries. The balanced cohort was projected using UMAP with a neighborhood size of 15 and a minimum distance of 0.1. Class boundaries were derived using a bandwidth factor of 1.1 and a threshold at the twentieth percentile of within class densities.

#### Disease family grouping

To examine how disease categories relate to one another at a broader level, all affected participants, excluding the unaffected group, were pooled into a single cohort. Within this pooled cohort, diagnoses exceeding ten times the size of the smallest diagnosis were downsampled to this cap, reducing the disproportionate influence of the largest individual diagnoses while retaining the internal heterogeneity of families composed of several smaller diagnoses. This balanced cohort was projected into a single shared UMAP embedding using the same neighbourhood size and minimum distance as above, and boundaries were drawn for each disease family by grouping the diagnoses comprising that family, using a bandwidth factor of 1.5 and a threshold at the twentieth percentile of within group densities.

#### Diagnosis grouping within family

For each disease family, participants belonging to that family were further examined in isolation. Within each family, an independent UMAP embedding was fitted using only that family’s participants, rather than reusing the shared embedding described above, so that each family occupies its own distinct projection. Boundaries within each family panel were drawn per individual diagnosis rather than per family, using the same neighbourhood size, minimum distance, bandwidth factor, and percentile threshold as the family level analysis.

#### Individual subject

In addition to the cohort level figures described above, a UMAP panel was generated as part of each individual subject’s report, contextualizing that subject’s position relative to their predicted diagnosis and a comparison diagnosis. The comparison diagnosis was taken as the model’s second ranked predicted diagnosis where more than one diagnosis was predicted, or could instead be manually specified to compare the subject against a particular clinically relevant alternative. The cohort was restricted to only those participants carrying either the subject’s top predicted diagnosis or this comparison diagnosis, focusing the panel on the relationship between these two categories rather than the full cohort. For each of the two diagnostic groups, an outer boundary was estimated using the same Gaussian kernel density approach described above, with a bandwidth factor of 1.5 and a threshold at the 20th percentile of within-group densities, consistent with the disease family and within family analyses. The subject’s own morphological deviation profile was projected into this same embedding space, using the fitted reducer where a new embedding had been generated and marked distinctly on the panel relative to the two group boundaries.

### Method 4: Predictive modeling

To evaluate the diagnostic discriminative power of individual and combined morphological descriptors, we used a supervised learning framework for multiclass classification and binary one-vs-rest classification. Separate independent models were trained and evaluated for each feature set to enable direct comparison of predictive performance across different morphological descriptors.

### Model Training and Evaluation

Predictive performance was estimated using group-aware cross-validation to prevent leakage from repeated measurements. Samples were assigned to 10 folds using a stratified group-aware split, preserving class proportions while ensuring that all repeated measurements from the same subject remained within a single fold. To mitigate class imbalance, diagnosis level downsampling was applied within each training fold only, leaving the held out fold unchanged. For each training split, class counts were calculated across the diagnosis labels and the smallest class count was used as the reference. An upper cap was then defined as this reference count multiplied by a fixed multiplier, set to 1 by default. Any diagnostic class with more samples than this cap was randomly downsampled to the cap before model fitting, whereas classes at or below the cap were retained in full. This procedure was applied uniformly across diagnostic classes, including the positive class in binary analyses. Models were implemented as scikit-learn pipelines comprising feature standardization followed by a linear Ridge classifier. For multi-feature sets (surface, volume and all features combined), a Random Forest classifier was also used to evaluate whether additional model complexity improved performance. For each feature set, the reported results correspond to the better-performing classifier (Ridge or Random Forest). Models were configured for class-balanced learning. Fitting and prediction were performed independently within each fold, and out-of-fold (OOF) predictions and class probabilities were aggregated to yield an unbiased estimate for each sample. Statistical significance of predictive performance was assessed using a non-parametric permutation test (10,000 permutations).

### Method 4a: Binary (One vs Rest) Classification

In the binary setting, classification was performed in a one-vs-rest manner, either for a single predefined diagnosis or iteratively across all diagnostic groups. Labels were binarized within each run and performance metrics were computed for the positive diagnosis of interest. Binary performance was assessed using balanced accuracy, precision, recall, F1-score, ROC AUC, and average precision, computed from OOF predictions and probabilities. Statistical significance was estimated using a permutation test with 10,000 permutations, from which the reported p-values were derived.

### Method 4b: Multiclass Classification

In the multiclass setting, models were trained to jointly discriminate between all diagnostic categories. Performance was summarized using overall accuracy, balanced accuracy, and macro-averaged precision, recall and F1-score, which weight diagnostic classes equally. Discrimination was further quantified using macro-averaged one-vs-rest ROC AUC and macro-averaged precision–recall (average precision) computed from calibrated probability outputs. Confusion matrices accumulated across folds were used to characterize class-specific error patterns.

### Method 5: Radar Plotting of Morphological Profiles

Radar plots were generated to visualize disease-specific morphological deviation profiles relative to healthy controls. For each diagnosis, cases were individually matched 1:1 to healthy controls by sex and nearest age using a greedy algorithm that prevented control reuse within each disease comparison. Morphological descriptors were standardized as

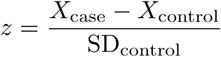

where SD_control_ was computed across the matched control group for that diagnosis. Analyses were stratified by sex, and male and female trajectories were displayed using distinct line styles (dotted and dashed, respectively) with shaded polygons. Per-measure significance was assessed using paired t-tests (when differences were approximately normally distributed) or Wilcoxon signed-rank tests otherwise; significant comparisons were indicated by appending markers to the corresponding axis label (* for males, *†* for females when p *<* 0.05). All radar plots were centred at zero (matched-control mean) and radial axes were clipped to ±3 SD to improve interpretability.

### Method 6: Individual Profile Report

### Individual level inference

To interrogate the reference space at the patient level, we generated an individual diagnostic report for each target subject using a hold-out inference procedure. Prior to training, every record sharing the target subject’s stratification identifier was removed from the cohort, ensuring that no measurement originating from the same individual, including any repeated visits, contributed to the models used to classify that subject. The remaining participants formed the training set. Inference proceeded hierarchically across three nested levels of increasing diagnostic specificity. The first level separated clinical status (healthy versus affected), the second assigned a broad disease family, and the third assigned a specific diagnosis. At each level a classifier was trained on the current training set and applied to the held out subject, yielding a predicted label and its associated posterior probability. After each level other than the last, the training set was restricted to those participants whose label at that level matched the prediction made for the target subject, so that the next and more specific classifier was fitted only on the relevant subpopulation. At the diagnosis level the three most probable diagnoses and their associated probabilities were retained rather than only the highest-ranked label, allowing the report to represent diagnostic uncertainty and possible comorbidity.

#### Model training

Each classifier used a random forest algorithm taking the complete morphological deviation profile, age and sex as input. Class imbalance across diagnostic categories was corrected by applying the same technique outlined in Method 4. All models were fitted with a fixed random seed for reproducibility, and a fresh unfitted copy of the pipeline was cloned at every level to prevent parameter carry over between levels.

#### Report composition

For each subject the pipeline assembled a single report combining the model inference with the underlying morphological deviation profile. The report presented the subject’s standardized descriptor values as a ranked profile, the predicted probabilities at each hierarchical level, and, where ground truth labels were available, an indication of whether each prediction was correct. For participants with a recorded comorbidity, a prediction was treated as correct when both the primary diagnosis and the comorbid condition appeared among the two most probable diagnoses. Two further panels situated the subject relative to the predicted diagnosis group. A radial profile (Method 5) compared the subject’s morphological deviation profile against the mean profile of the predicted diagnosis group. A UMAP embedding (Method 3) of the two most probable diagnosis groups placed the subject within the morphology reference space. The target subject was projected into the same space by out of sample transformation and marked as a distinct point. Participants carrying both of the two displayed diagnoses as comorbid conditions were highlighted.

#### Longitudinal analysis

For participants with repeated visits, the report was extended with a longitudinal summary. The predicted probability of each candidate diagnosis was tracked across visits and fitted with a low order polynomial to indicate temporal trends. In parallel, the subject’s position at each visit was projected into a shared UMAP embedding of the candidate diagnosis groups, and consecutive visits were connected by directional arrows to visualize the subject’s trajectory through the morphological reference space over time.

### Method 7: Cox Proportional Hazards modeling for Longitudinal Risk Assessment

To assess disease progression risk across different clinical trajectories, we employed Cox proportional hazards (Cox PH) models using the lifelines package (https://github.com/CamDavidsonPilon/lifelines/). Three distinct conversion scenarios were modeled: HC to AD, HC to MCI, and MCI to AD, with time-to-event defined as years since baseline for HC-initiated transitions and years since MCI diagnosis for the MCI-to-AD transition. For each scenario, survival analysis datasets were constructed in a time-static framework, where baseline observations (first visit without the target event) were extracted for each subject, and event times were determined from the first occurrence of diagnostic conversion. The models incorporated age, sex, and the target morphological descriptor z-scores (normalized relative to the HC reference distribution) as covariates. To ensure independent evaluation of the visualised longitudinal case, models were trained on participants excluding both the progressive converter and stable HC individuals shown in the longitudinal example. Sex was included as a stratification variable to account for baseline hazard differences between males and females while maintaining proportional hazards assumptions for the remaining covariates. To prevent zero-duration events, a small constant (0.01 years) was added to all survival times. For longitudinal risk trajectory visualization, we selected representative participants from four clinically relevant groups: full converters (participants progressing from HC through MCI to AD), stable HC (participants remaining cognitively normal throughout follow-up), stable MCI (participants entering the study with MCI and retain this status without conversion to AD), and incidental MCI (participants with baseline HC diagnosis who convert to MCI but do not convert to AD). Individual survival probabilities were computed at each visit time point using a conditional approach, treating each subsequent visit as a new baseline for risk assessment over an 8-year prediction horizon. This visit-conditional framework allowed tracking of temporal changes in conversion probability throughout each subject’s clinical trajectory. For participants transitioning between diagnostic states, the model applied at each visit depended on the participant’s current diagnosis. Individuals who converted to MCI were evaluated using the HC-to-AD model before MCI conversion and the MCI-to-AD model thereafter, whereas participants who never converted were evaluated exclusively using the HC-to-AD model. Event probabilities were computed as 1 minus the survival function at visit time plus the specified horizon, representing the probability of experiencing the target conversion event within the prediction window.

## Data Availability

All data used in the manuscript are openly available online.

## Declarations

### Funding

C. Dalby was supported by a PhD grant by the Medical Research Council (MRC) as part of the Precision Medicine Doctoral Training Programme (MRC, grant number: MR/W006804/1). A. Dibble was supported by a PhD grant from the Scottish Graduate School of Social Science, Doctoral Training Partnership (SGSSS-DTP), on behalf of the Economic and Social Research Council (ESRC, grant number: ES/P000681/1). A.Fracasso was supported by a grant from the Biotechnology and Biological Sciences Research Council (BBSRC, grant number: BB/S006605/1) and the Bial Foundation (Bial Foundation Grants Programme; Grant id: A-29315, number: 203/2020, grant edition: G-15516). This research has been conducted using the UK Biobank Resource under Application 17689.

### Author contributions (CRediT): Connor Dalby

Conceptualization, Methodology, Software, Validation, Formal Analysis, Investigation, Data Curation, Writing - Original Draft, Writing - Review & Editing, Visualization. **Austin Dibble:** Conceptualization, Methodology, Validation, Formal Analysis, Writing - Review & Editing, Data Curation, Visualization. **Damiano Ferrari:** Conceptualization, Methodology, Validation, Formal Analysis, **Sergio Benini:** Conceptualization, Writing - Review & Editing, Supervision. **Alessio Fracasso:** Conceptualization, Methodology, Writing - Review & Editing, Supervision. **Donald M. Lyall:** Methodology, Resources, Writing - Review & Editing, Data Curation. **Terry Quinn:** Writing - Review & Editing, **Lars Muckli:** Writing - Review & Editing, Funding Acquisition, **Monika Harvey:** Writing - Review & Editing, Funding Acquisition, **Michele Svanera:** Conceptualization, Investigation, Methodology, Software, Writing - Original Draft, Writing - Review & Editing, Data Curation, Visualization, Resources, Supervision, Funding Acquisition, Project Administration.

## Acknowledgements

Some of the data used in the preparation of this article were obtained from the Frontotemporal Lobar Degeneration Neuroimaging Initiative (FTLDNI). For up-to-date information on participation and protocol, see http://memory.ucsf.edu/research/studies/nifd.

Data were provided [in part] by the Human Connectome Project, WU-Minn Consortium (Principal Investigators: David Van Essen and Kamil Ugurbil; 1U54MH091657) funded by the 16 NIH Institutes and Centers that support the NIH Blueprint for Neuroscience Research; and by the McDonnell Center for Systems Neuroscience at Washington University.’

Data were provided [in part] by OASIS-3: Longitudinal Multimodal Neuroimaging: (Principal Investigators: T. Benzinger, D. Marcus, J. Morris); NIH P30 AG066444, P50 AG00561, P30 NS09857781, P01 AG026276, P01 AG003991, R01 AG043434, UL1 TR000448, R01 EB009352. AV-45 doses were provided by Avid Radiopharmaceuticals, a wholly owned subsidiary of Eli Lilly.

Data collection and sharing for this project was funded by the Alzheimer’s Disease Neuroimaging Initiative (ADNI) (National Institutes of Health Grant U01 AG024904) and DOD ADNI (Department of Defense award number W81XWH-12-2-0012). ADNI is funded by the National Institute on Aging, the National Institute of Biomedical Imaging and Bioengineering, and through generous contributions from the following: AbbVie, Alzheimer’s Association; Alzheimer’s Drug Discovery Foundation; Araclon Biotech; BioClinica, Inc.; Biogen; Bristol-Myers Squibb Company; CereSpir, Inc.; Cogstate; Eisai Inc.; Elan Pharmaceuticals, Inc.; Eli Lilly and Company; EuroImmun; F. Hoffmann-La Roche Ltd and its affiliated company Genentech, Inc.; Fujirebio; GE Healthcare; IXICO Ltd.; Janssen Alzheimer Immunotherapy Research & Development, LLC.; Johnson & Johnson Pharmaceutical Research & Development LLC.; Lumosity; Lundbeck; Merck & Co., Inc.; Meso Scale Diagnostics, LLC.; NeuroRx Research; Neurotrack Technologies; Novartis Pharmaceuticals Corporation; Pfizer Inc.; Piramal Imaging; Servier; Takeda Pharmaceutical Company; and Transition Therapeutics. The Canadian Institutes of Health Research is providing funds to support ADNI clinical sites in Canada. Private sector contributions are facilitated by the Foundation for the National Institutes of Health (www.fnih.org). The grantee organization is the Northern California Institute for Research and Education, and the study is coordinated by the Alzheimer’s Therapeutic Research Institute at the University of Southern California. ADNI data are disseminated by the Laboratory for Neuro Imaging at the University of Southern California

Data used in the preparation of this article was obtained on 2025-08-18 from the Parkinson’s Progression Markers Initiative (PPMI) database (www.ppmi-info.org/access-dataspecimens/download-data), RRID:SCR 006431. For up-to-date information on the study, visit www.ppmi-info.org. PPMI - a public-private partnership - is funded by the Michael J. Fox Foundation for Parkinson’s Research, and funding partners; including [insert full list of all PPMI funding partners found on the PPMI Website.

Data collection and dissemination of the data presented in this manuscript was supported by the ALLFTD Consortium (U19: AG063911, funded by the National Institute on Aging and the National Institute of Neurological Diseases and Stroke) and the former ARTFL & LEFFTDS Consortia (ARTFL: U54 NS092089, funded by the National Institute of Neurological Diseases and Stroke and National Center for Advancing Translational Sciences; LEFFTDS: U01 AG045390, funded by the National Institute on Aging and the National Institute of Neurological Diseases and Stroke). The manuscript has been reviewed by the ALLFTD Executive Committee for scientific content. The authors acknowledge the invaluable contributions of the study participants and families as well as the assistance of the support staffs at each of the participating sites.

Data collection and sharing for this project was funded by the Frontotemporal Lobar Degeneration Neuroimaging Initiative (National Institutes of Health Grant R01 AG032306). The study is coordinated through the University of California, San Francisco, Memory and Aging Center. FTLDNI data are disseminated by the Laboratory for Neuro Imaging at the University of Southern California.

Data used in preparation of this article were shared by the Mayo Clinic Study of Aging (MCSA). The MCSA is funded by the following sources: NIH U01 AG006786, R01 AG034676, R37 AG011378, R01 AG041851, R01 NS097495, R01 AG056366, R01 AG068206, P30 AG062677, GHR Foundation, Elsie and Marvin Dekelboum Family Foundation, Liston Award, Schuler Foundation, Alexander Foundation, Mayo Foundation for Medical Education and Research.

Data were provided in part by OASIS-3: Longitudinal Multimodal Neuroimaging: Principal Investigators: T. Benzinger, D. Marcus, J. Morris; NIH P30 AG066444, P50 AG00561, P30 NS09857781, P01 AG026276, P01 AG003991, R01 AG043434, UL1 TR000448, R01 EB009352, P30 AG066444, AW00006993. AV-45 doses were provided by Avid Radiophar-maceuticals, a wholly owned subsidiary of Eli Lilly. AV-1451 doses were provided by Avid Radiopharmaceuticals, a wholly owned subsidiary of Eli Lilly.

Data used in the preparation of this article were obtained from the Adolescent Brain Cognitive Development (ABCD) Study (https://abcdstudy.org), held in the NIMH Data Archive (NDA). This is a multisite, longitudinal study designed to recruit more than 10,000 children aged 9–10 and follow them over 10 years into early adulthood. The ABCD Study® is supported by the National Institutes of Health and additional federal partners under award numbers U01DA041048, U01DA050989, U01DA051016, U01DA041022, U01DA051018, U01DA051037, U01DA050987, U01DA041174, U01DA041106, U01DA041117, U01DA041028, U01DA041134, U01DA050988, U01DA051039, U01DA041156, U01DA041025, U01DA041120, U01DA051038, U01DA041148, U01DA041093, U01DA041089, U24DA041123, U24DA041147. A full list of supporters is available at https://abcdstudy.org/federal-partners.html. A listing of participating sites and a complete listing of the study investigators can be found at https://abcdstudy.org/consortium members. ABCD Consortium investigators provided data but did not necessarily participate in the analysis or writing of this report. This manuscript reflects the views of the authors and may not reflect the opinions or views of the NIH or ABCD Consortium investigators. Data use in preperation of this article were obtained from the IXI dataset website

## Notes

### Competing Interest Statement

The authors have declared no competing interest.

### Author Declarations

Datasets included human MRI datasets only, all openly available to the research community. A full breakdown of the datasets and relevant citations are available under Manuscript Supplementary Files. All datasets used in this study had been de-identified by the respective data providers prior to our access and use. We did not have access to any personally identifiable information, such as names, addresses, or other direct identifiers, at any stage of the study. Participants were represented only by de-identified/anonymised subject identifiers assigned by the original dataset providers, with any linkage to personal information retained solely by the relevant data custodians.

