## Supplementary Material for "NeuroMorph: A Unified Morphological Reference Space for Cross-Disease Brain Profiling"

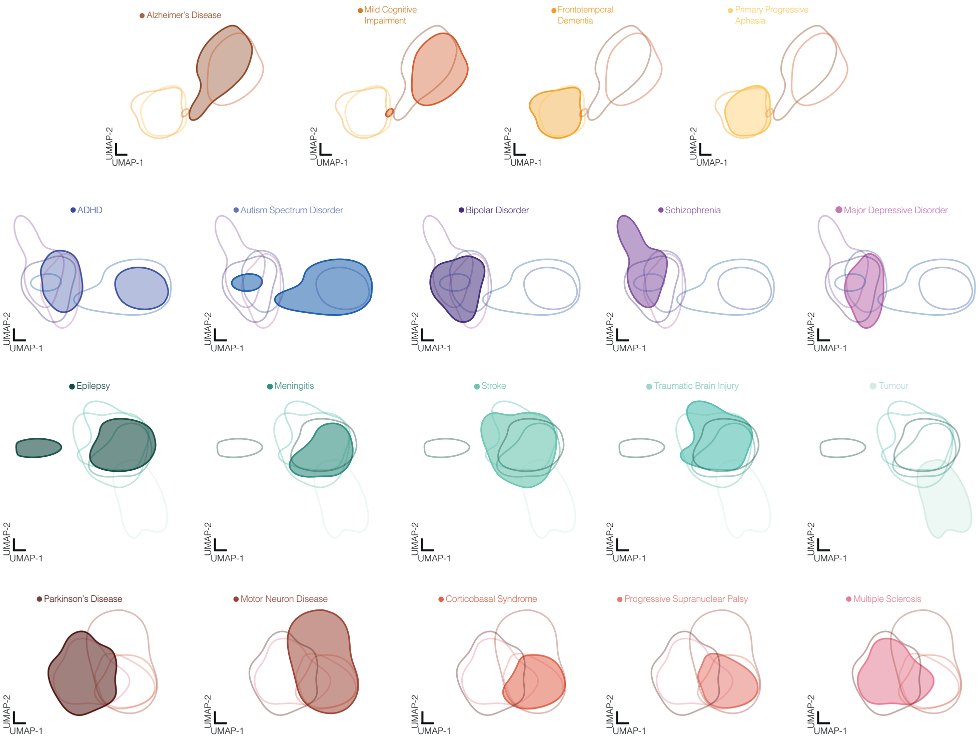


**Supplementary Fig 1**: **Diagnosis Levels UMAP Projections** UMAP projection of the shared morphological deviation space of the target condition in the Dementias (orange), Parkinsonism and Motor (red), Acquired Neurological (teal), and Psychiatric and Neurodevelopmental (blue) families. These projections are used for visualization only.

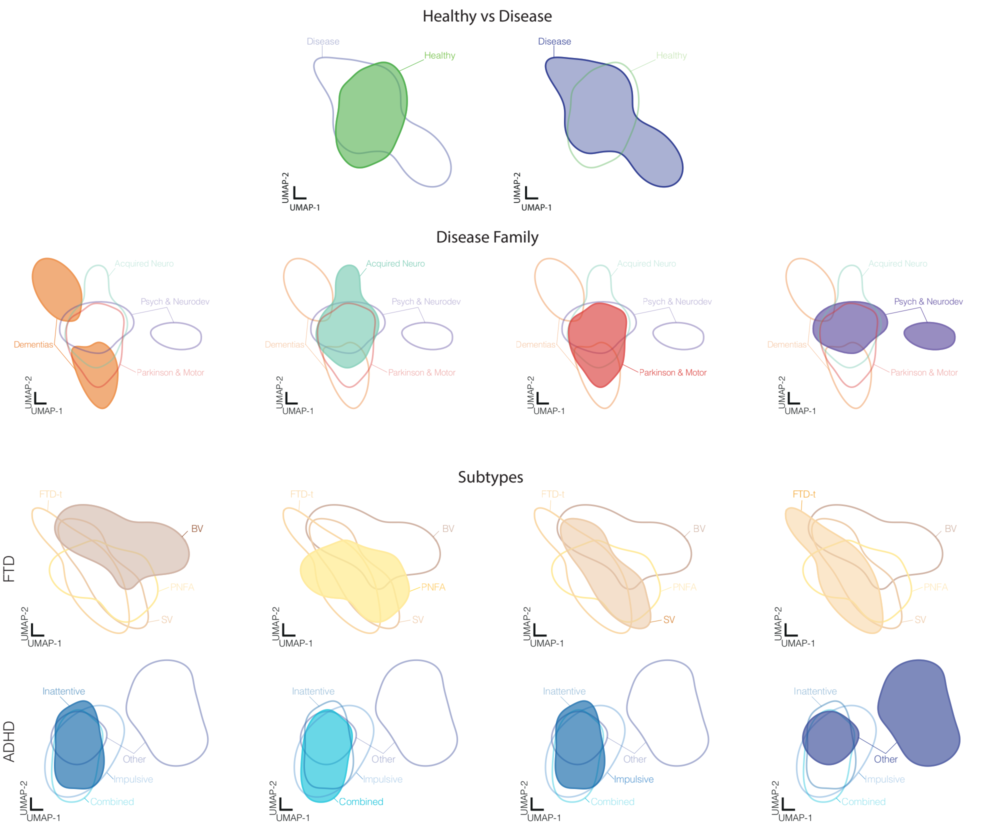


**Supplementary Fig 2**: **Clinical, Disease Family and Subtype Levels UMAP Projections** UMAP projection of the shared morphological deviation space of the target condition for the healthy and disease groups (row I), disease family level (row II), plus subtypes for FTD (row III) and ADHD (IV). These projections are used for visualization only.

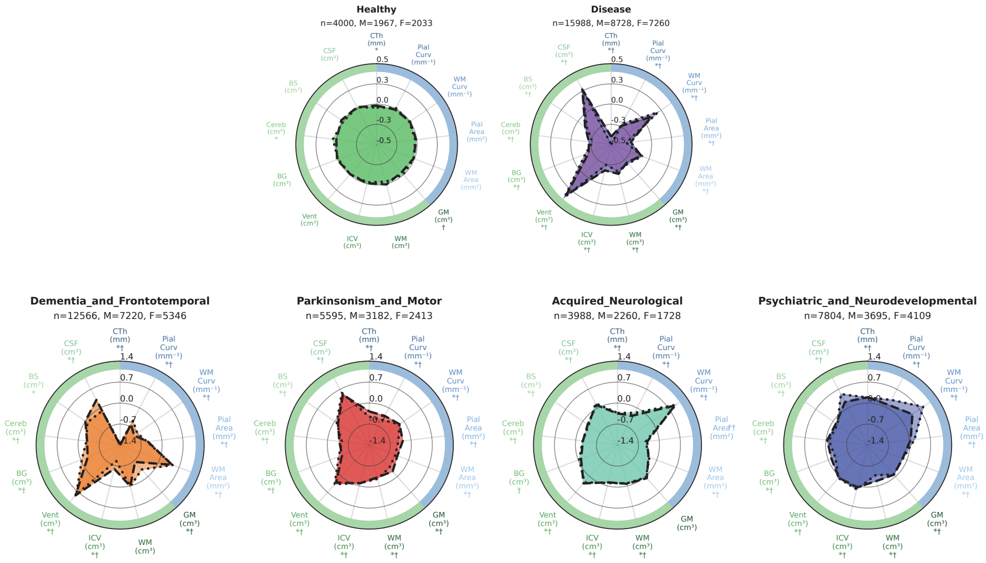


**Supplementary Fig 3**: **Higher Levels Radar Profiles** Morphological feature profile of the target condition for healthy and disease groups (row I) and each disease family (row II) level, represented as a radar plot. The centre circle represents the age and sex controlled healthy cohort. Significant differences between male and female cohorts are denoted by * and †, respectively.

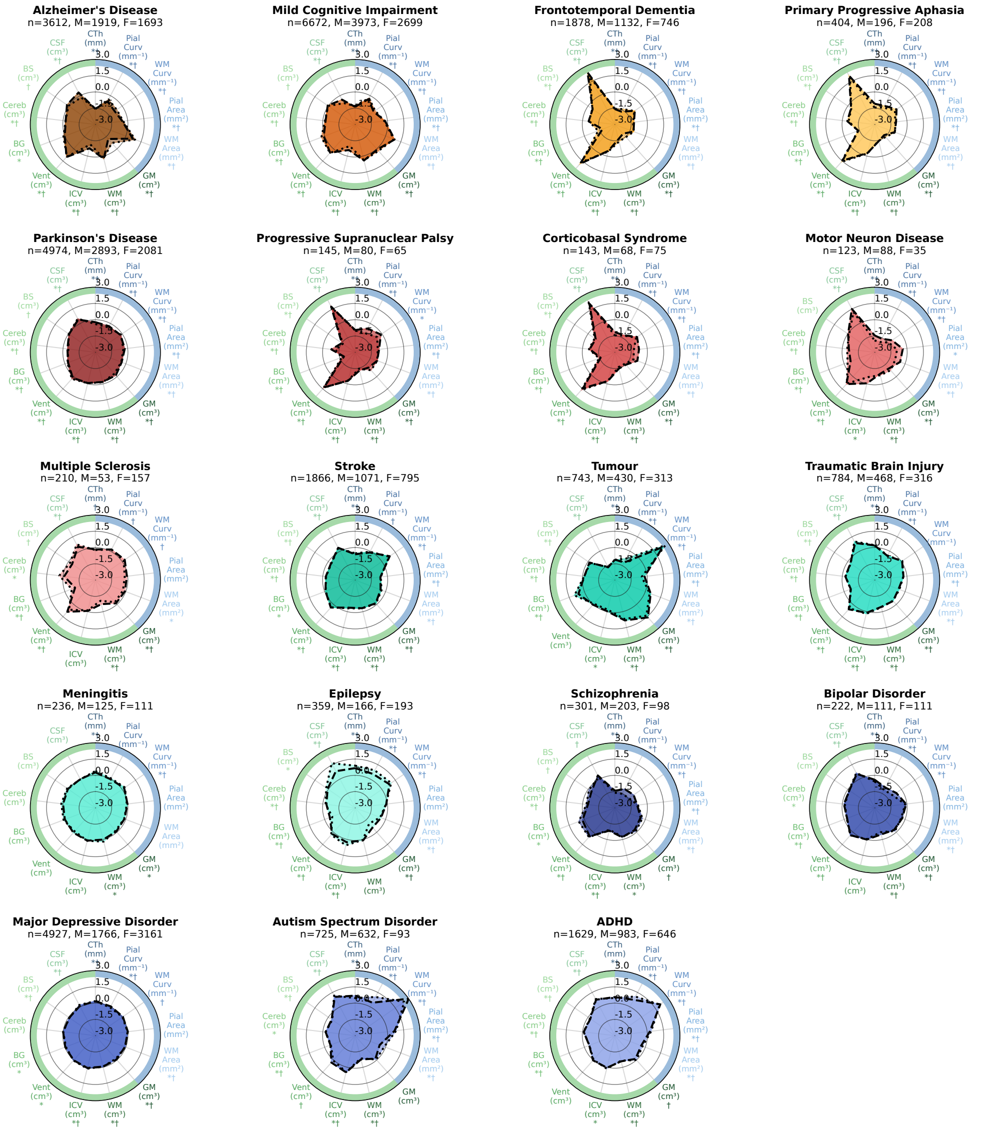


**Supplementary Fig 4**: **Diagnosis Levels Radar Profiles** Morphological feature profile of the target condition in the Dementias (orange), Parkinsonism and Motor (red), Acquired Neurological (teal), and Psychiatric and Neurodevelopmental (blue) families, represented as a radar plot. The centre circle represents the age and sex controlled healthy cohort. Significant differences between male and female cohorts are denoted by * and †, respectively.

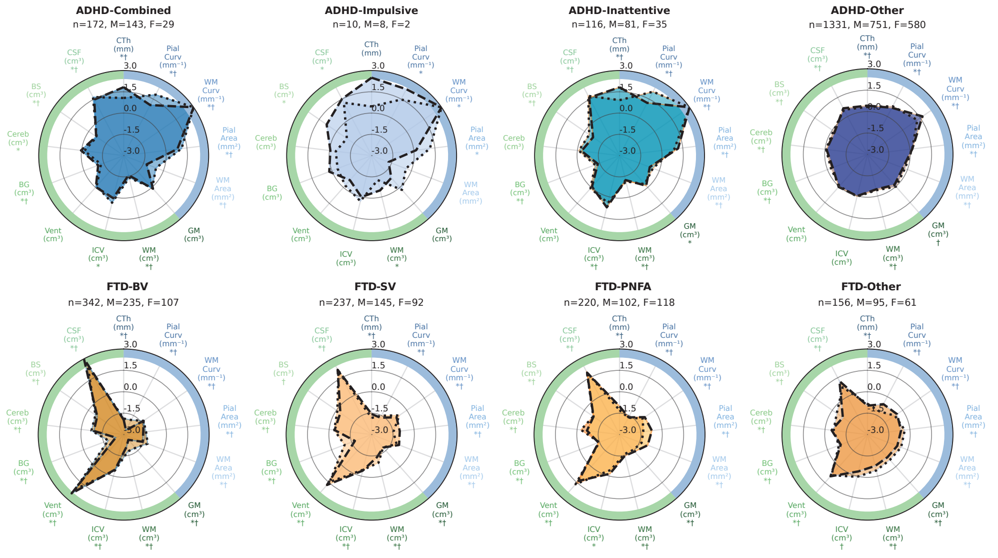


**Supplementary Fig 5**: **Subtype Levels Radar Profiles** Morphological feature profile of the target subtype in the ADHD (row I - blue) and Frontotemporal Dementia (row II - orange) diagnosis groups, represented as a radar plot. The centre circle represents the age and sex controlled healthy cohort. Significant differences between male and female cohorts are denoted by * and †, respectively.

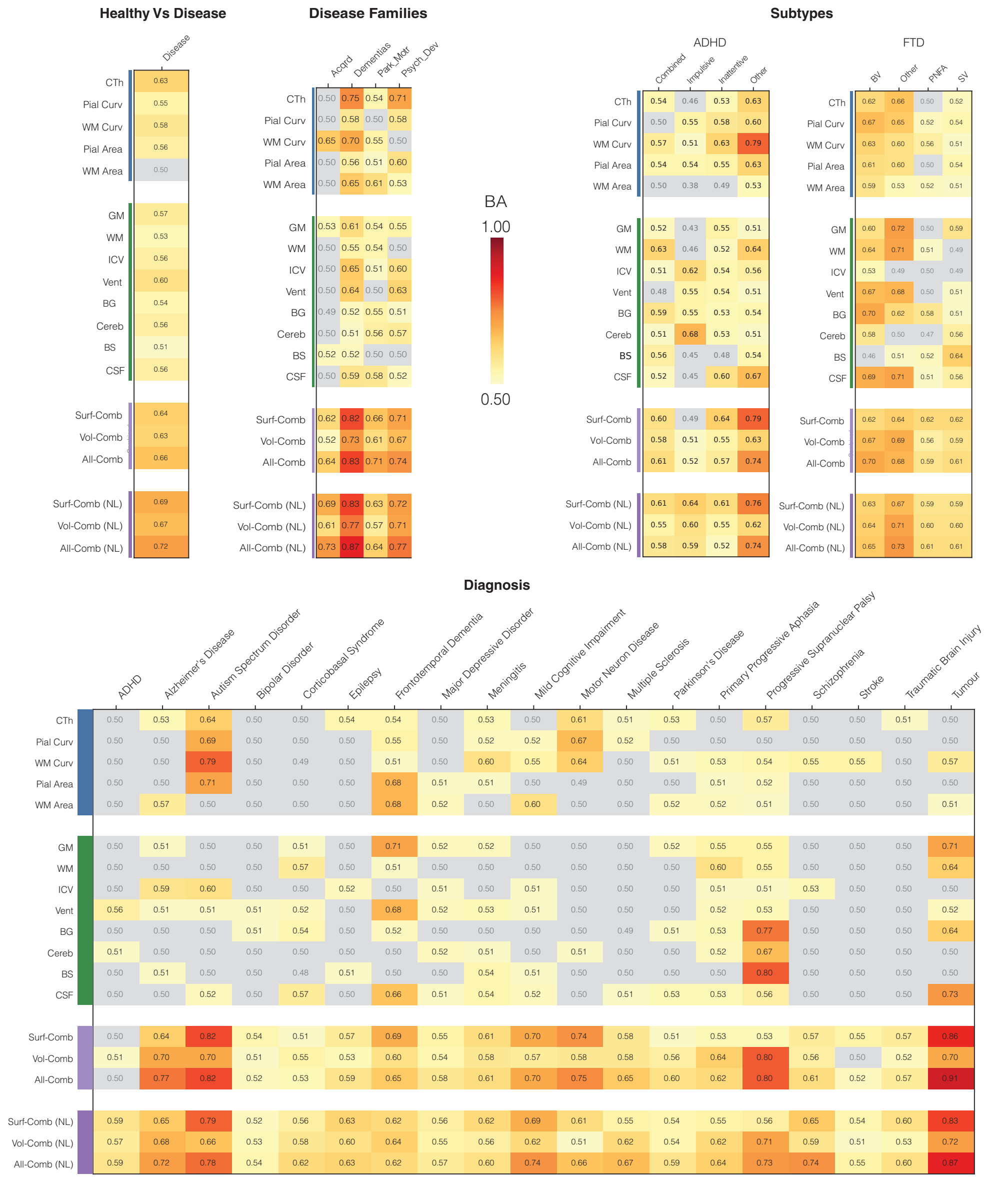


**Supplementary Fig 6**: **Balanced Accuracy Heatmaps** Descriptor-set performance heatmap in which each entry represents the balanced accuracy achieved by a given descriptor set when differentiating each condition from all others at the current level. Row labels in green and blue correspond to volumetric and surface-based descriptors evaluated in isolation, respectively, while purple labels denote combined descriptor sets incorporating all volumetric, surface-based, or morphological descriptors. All results generated using Ridge classifier models unless label has a ”NL” suffix which reflects the use of random forest classifiers. Balanced accuracy values approaching 1.0 indicate strong discrimination (red), while values approaching 0.5 reflect chance-level performance (white).

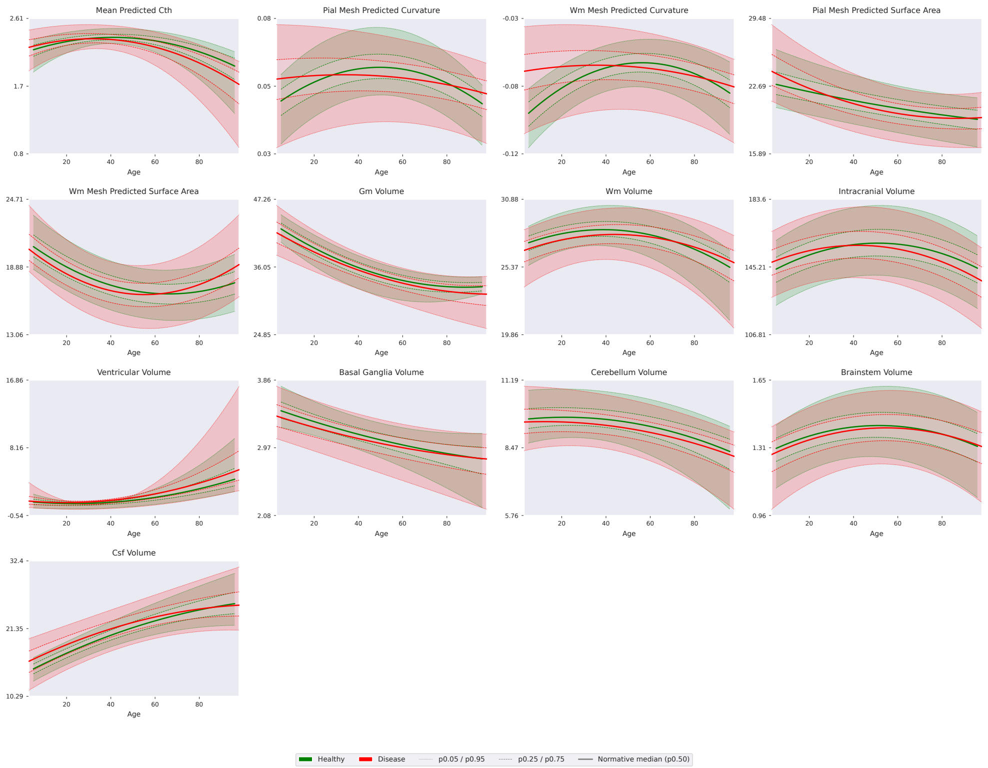


**Supplementary Fig 7**: **Brain Centile Charts For Healthy and Disease** Normative trajectories for the disease group (all conditions - red) relative to the healthy group (green) across the age range of the target cohort. Normative centiles are represented as follows: 0.5 (median) as a bold line, 0.25/0.75 as dashed lines, and 0.05/0.95 as dotted lines.

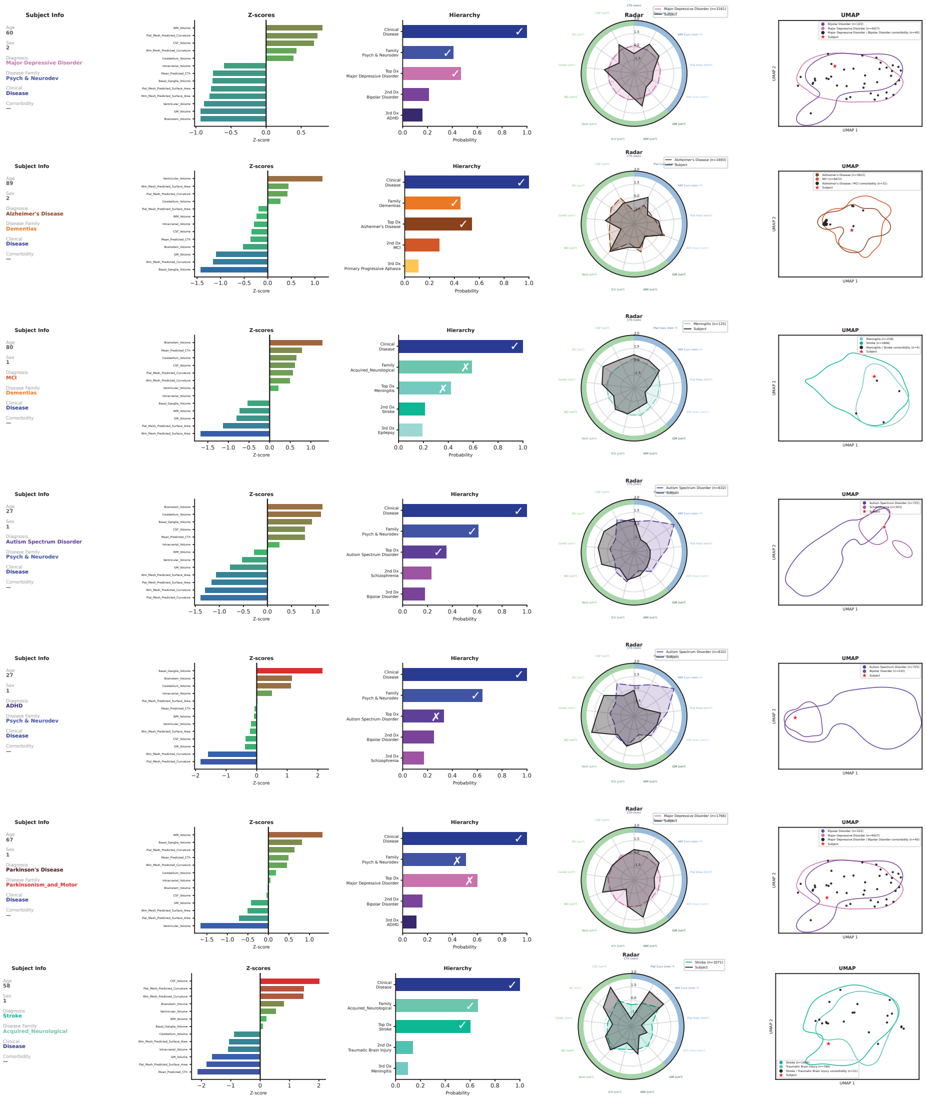

**Supplementary Fig 8**: **Profile Reports for Randomly Selected Individuals** Prediction reports for seven individuals sampled at random from the cohort. Reports were generated using Method 6 and follow the layout of the individual case examples in Figs 3 to 5. A tick denotes a morphology prediction that agrees with the clinical diagnosis and a cross denotes disagreement

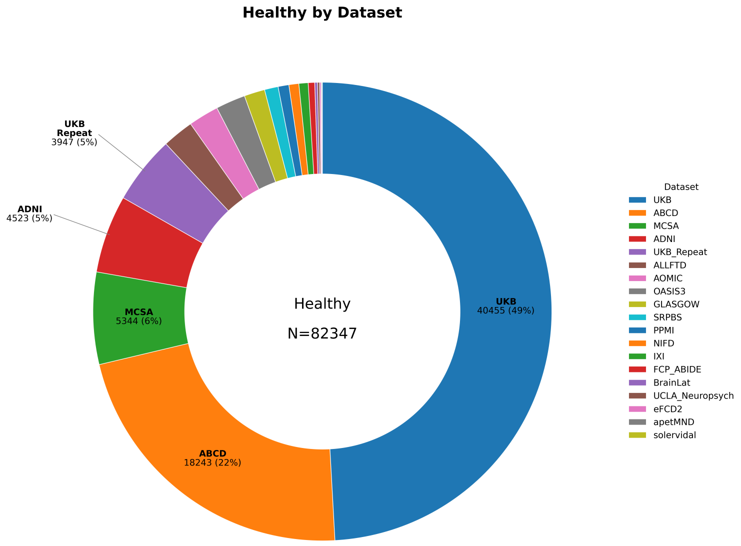


**Supplementary Fig 9**: **Composition of the Healthy Control Dataset** Breakdown of T1w MRI volume contributions from each dataset to the healthy group.

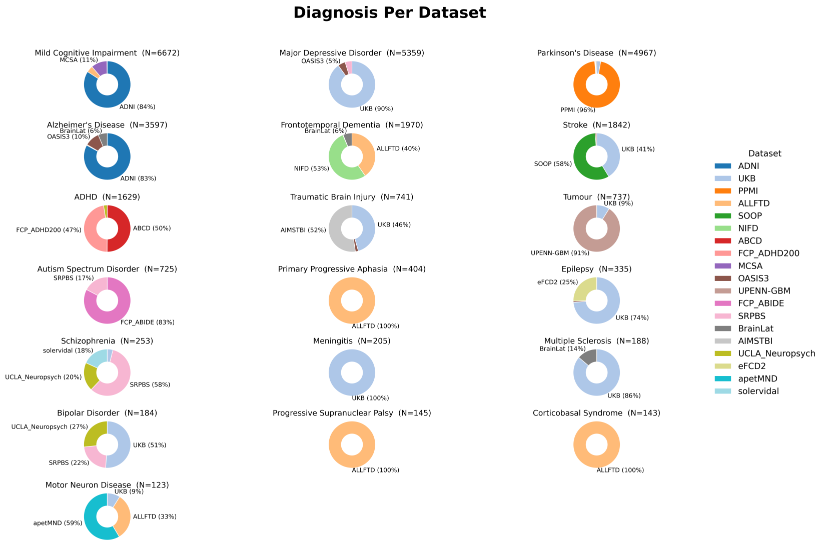


**Supplementary Fig 10**: **Diagnosis Per Dataset** Doughnut charts show the proportion of volumes contributed by each dataset for every diagnostic category.

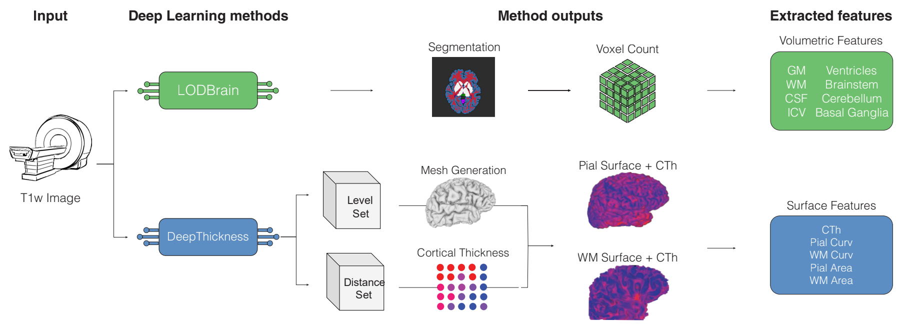


**Supplementary Fig 11**: **NeuroMorph Framework** A single T1w MR image is provided as input to the NeuroMorph framework, which leverages LOD-Brain+ and DeepThickness to generate an eight-mask segmentation volume, surface-level representation and distance set. Cortical surface reconstructions with CTh overlays are produced using marching cubes and linear interpolation. Voxel- and surface-based features are then extracted from the segmentation and reconstructed surfaces to form a complete morphological fingerprint.

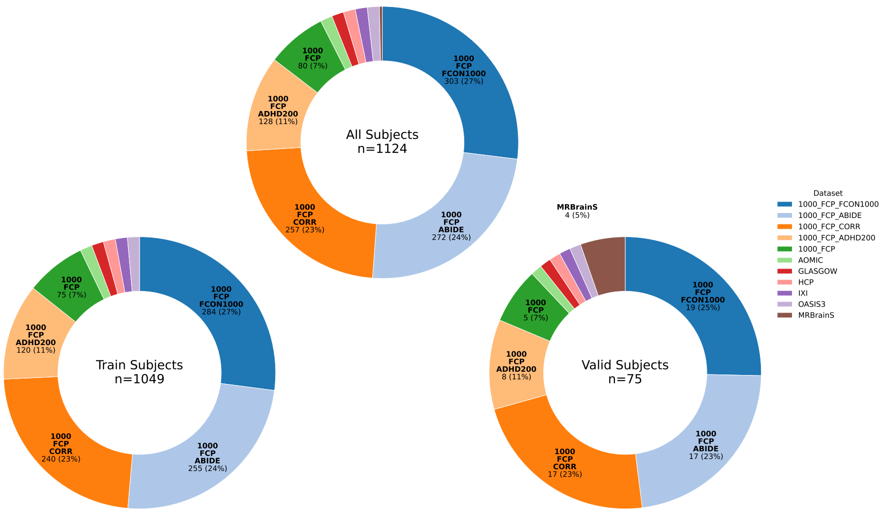


**Supplementary Fig 12**: **LOD-Brain+ Training Data** The datasets used for retraining the LOD-Brain model (top), split into train set (bottom-left) and validation set (bottom-right).

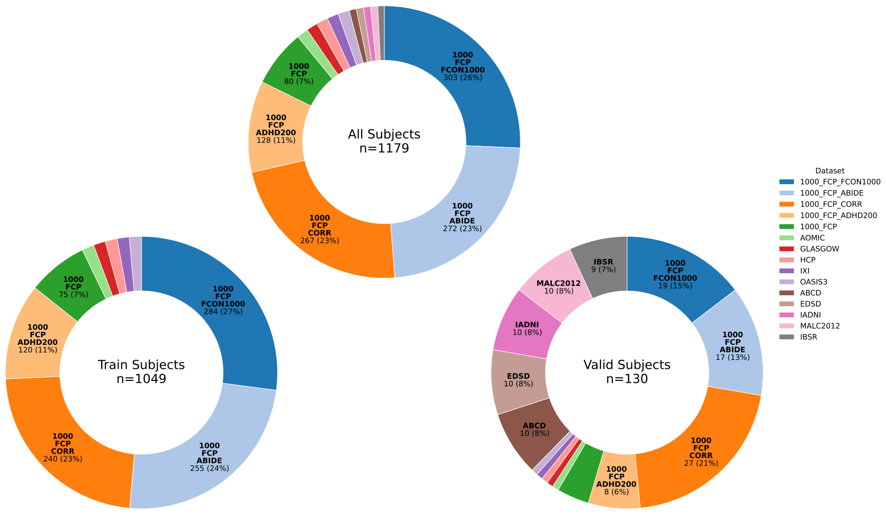


**Supplementary Fig 13**: **DeepThickness Training Data** The datasets used for training the DeepThickness model (top), split into train set (bottom-left) and validation set (bottom-right).

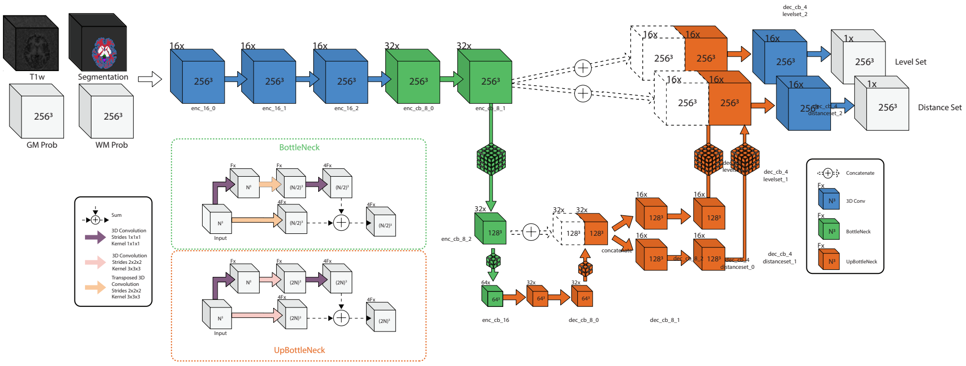


**Supplementary Fig 14**: **DeepThickness Model Architecture**

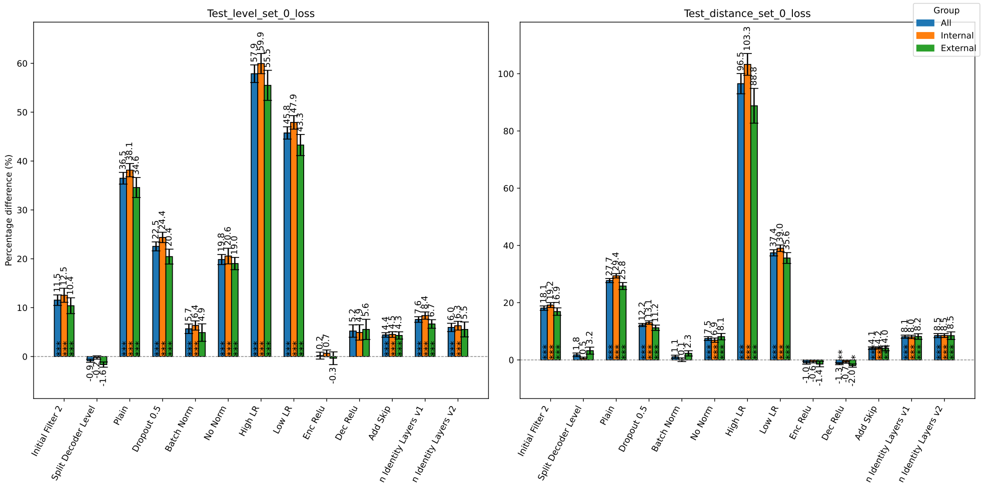


**Supplementary Fig 15**: **Ablation Study** Bar chart showing the impact of removing or modifying individual model components on performance, with bars indicating the resulting change relative to the final parameter choice.

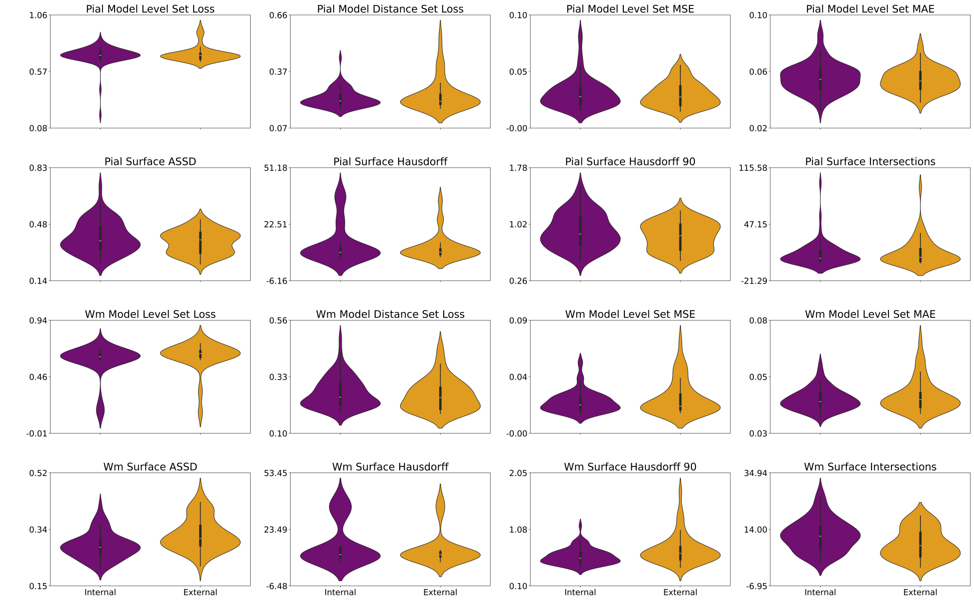


**Supplementary Fig 16**: **Model Performance Metrics** Violin plots show metric distributions for DeepThickness model volumetric outputs (row 1 = pial, row 3 = WM) and the corresponding cortical surface reconstructions (row 2 = pial, row 4 = WM) for the pial and WM surfaces, respectively. Metrics are reported on the validation set and stratified by volumes drawn from the training dataset (purple) or from a separate dataset not used during model training (orange).

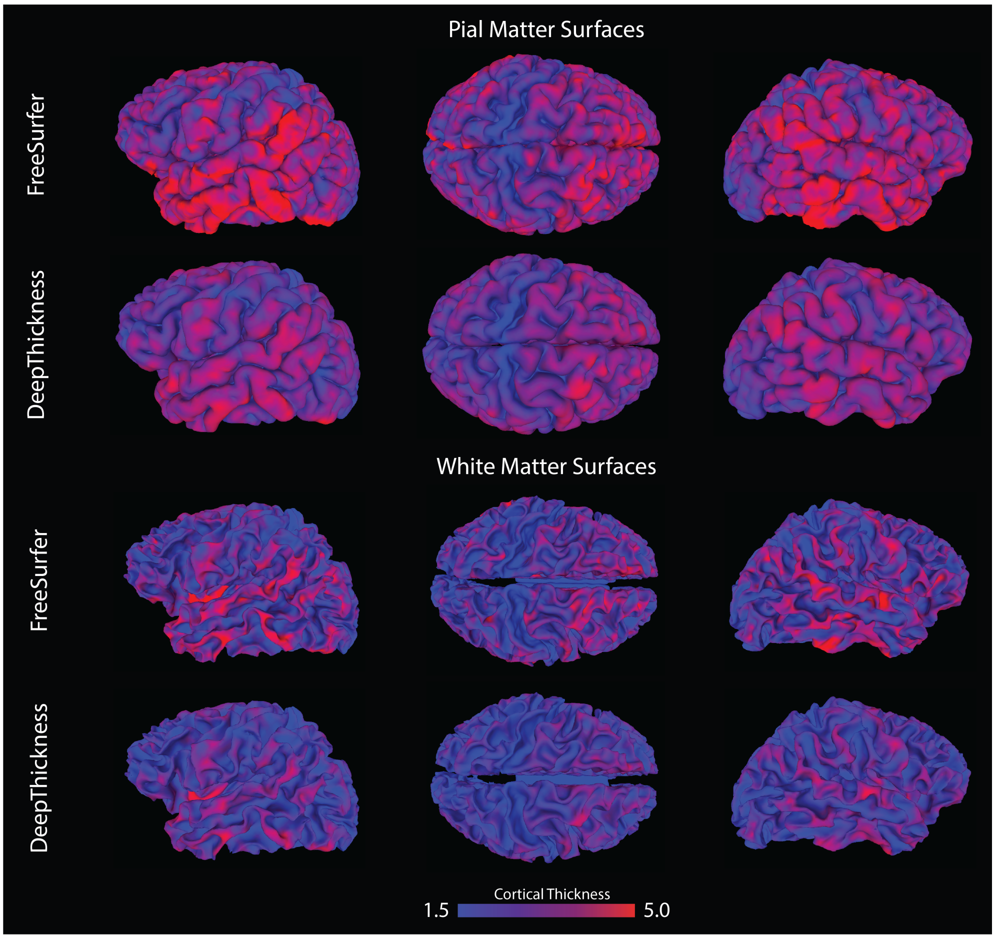


**Supplementary Fig 17**: **DeepThickness Generated Meshes** Examples of pial (panel 1) and WM (panel 2) mesh surfaces generated by FreeSurfer (top) compared with mesh surfaces generated by NeuroMorph (bottom). colors represent cortical thickness values either directly estimated by FreeSurfer or interpolated from the predicted distance set, respectively.

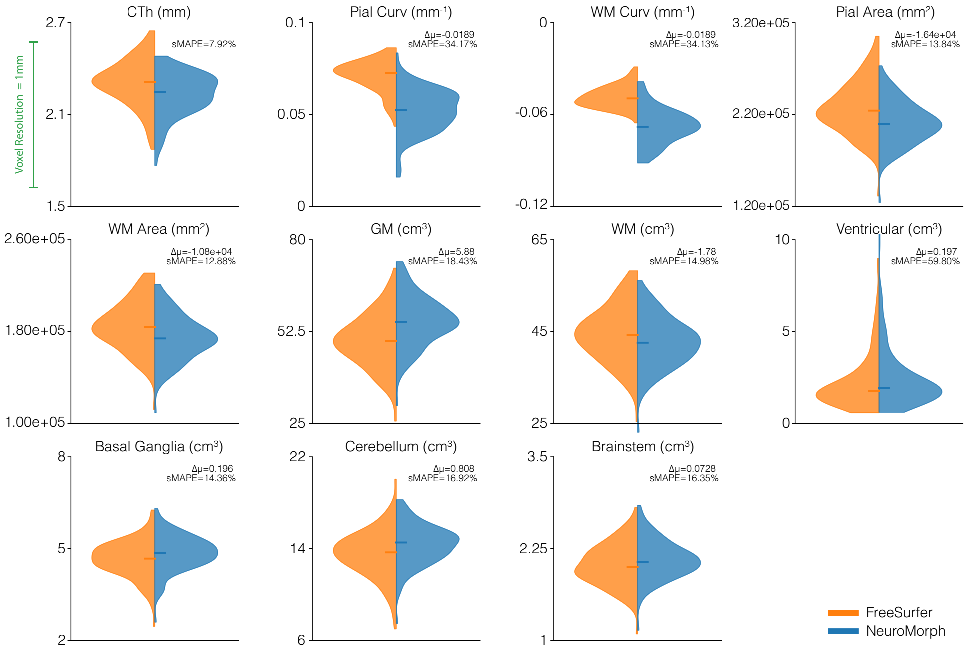


**Supplementary Fig 18**: **Quantitative Measure Evaluation: NeuroMorph Vs FreeSurfer** Morphological fingerprint features extracted from cortical segmentations and surface reconstructions generated by FreeSurfer (orange) and NeuroMorph (blue) are compared. Difference in FreeSurfer and NeuroMorph mean estimates ($\Delta\mu$) and Symmetric Mean Absolute Percentage Error (*sMAPE*) values are annotated for each measure. To ensure a fair comparison, an identical feature extraction procedure was applied to cortical segmentations and surface reconstructions generated by both methods.

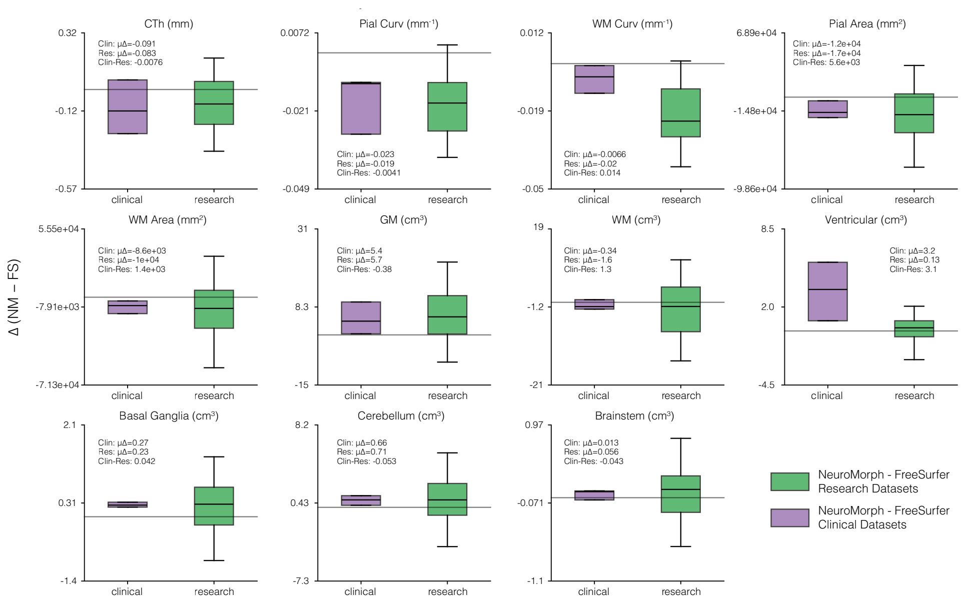


**Supplementary Fig 19**: **NeuroMorph vs FreeSurfer Split By Data Source** Per-feature differences between NeuroMorph and FreeSurfer (Δ) are shown for MRI volumes from clinical (purple) and research (green) datasets. Identical feature-extraction procedures were applied to outputs from both methods to enable a direct comparison.

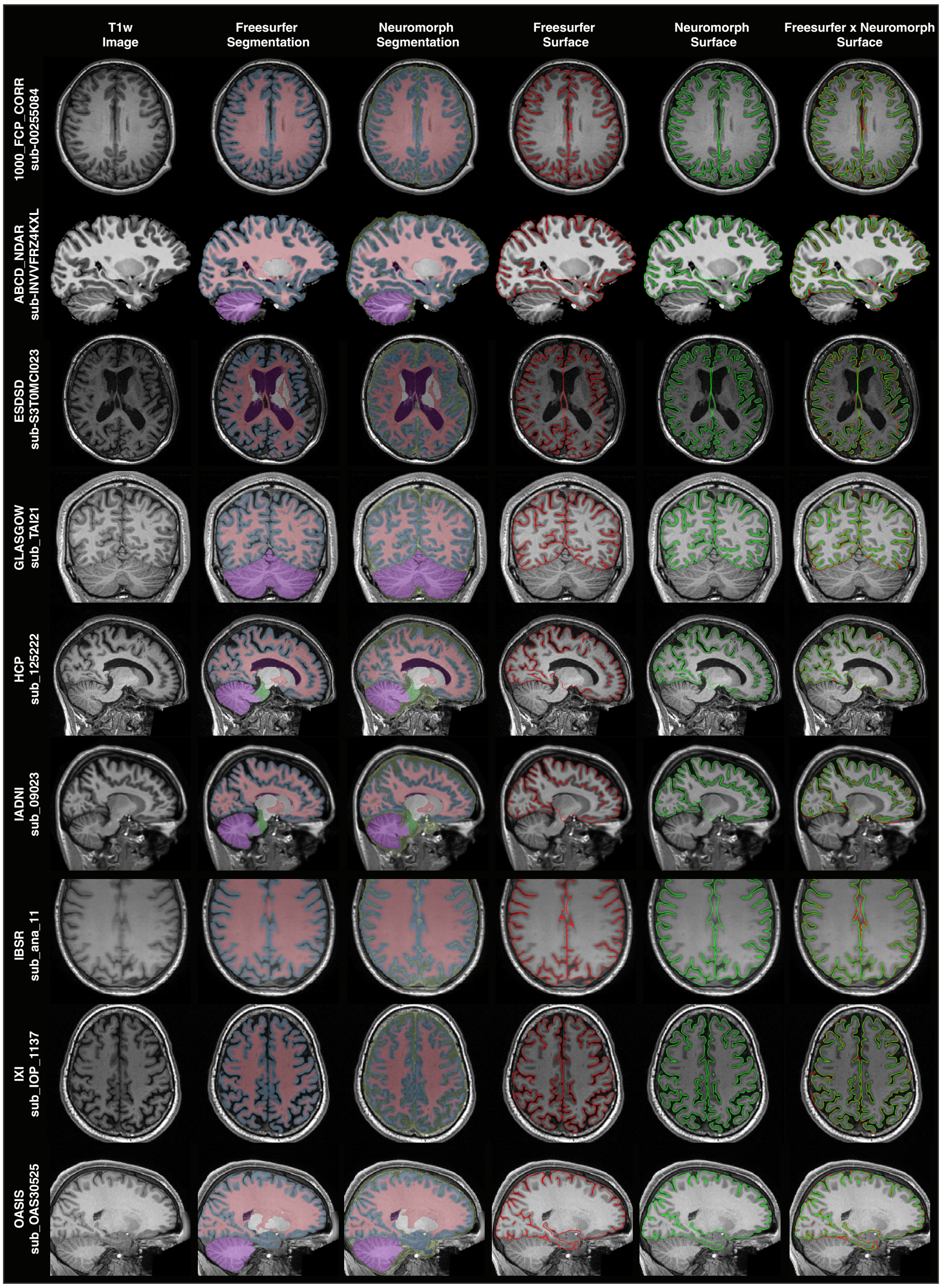


**Supplementary Fig 20**: **Voxel Space: NeuroMorph vs FreeSurfer Comparison** Single subject T1w MR images from nine independent datasets are shown with FreeSurfer segmentation (column 2), NeuroMorph segmentation (column 3), FreeSurfer pial surface reconstruction (red - column 4), NeuroMorph pial surface reconstruction (green - column 5) and overlaid FreeSurfer and NeuroMorph pial surfaces (column 6). Green outline in NeuroMorph segmentation marks CSF mask not generated by FreeSurfer.

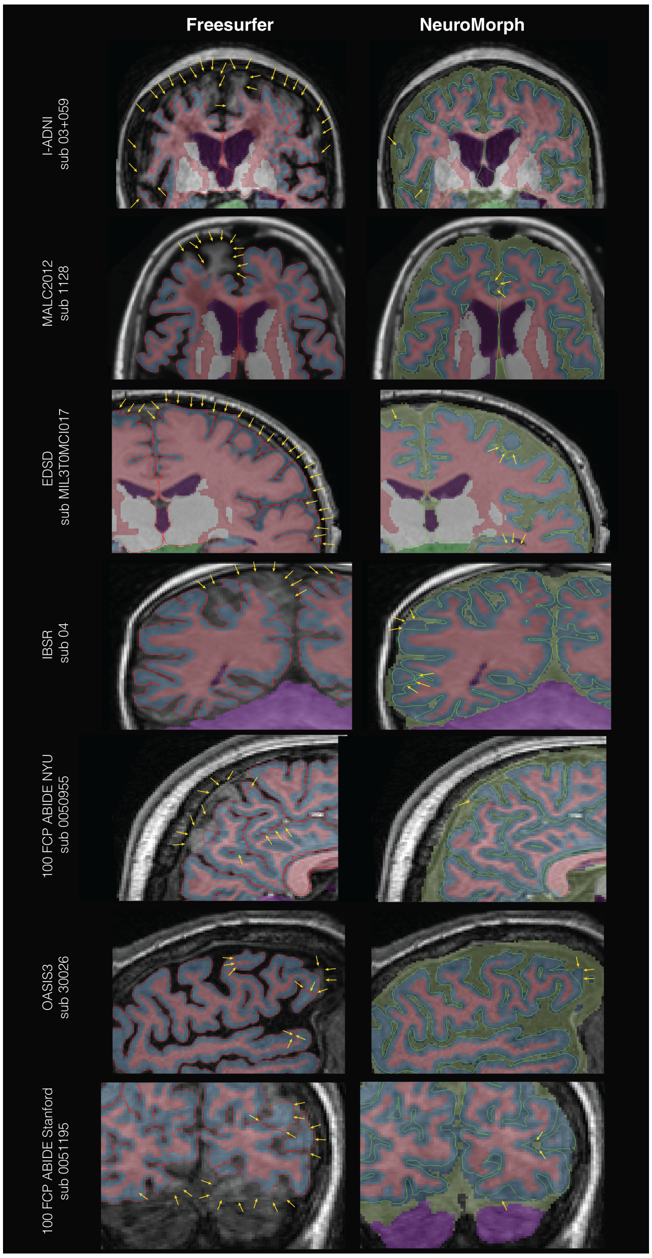


**Supplementary Fig 21**: **Error Comparison: FreeSurfer vs NeuroMorph** T1w MR images from seven individual participants drawn from different datasets were presented with cortical segmentation and surface reconstruction overlays generated by FreeSurfer (column 1) and NeuroMorph (column 2). Yellow arrows indicate regions of segmentation and/or surface reconstruction error. Green outline in NeuroMorph segmentation marks CSF mask not generated by FreeSurfer.

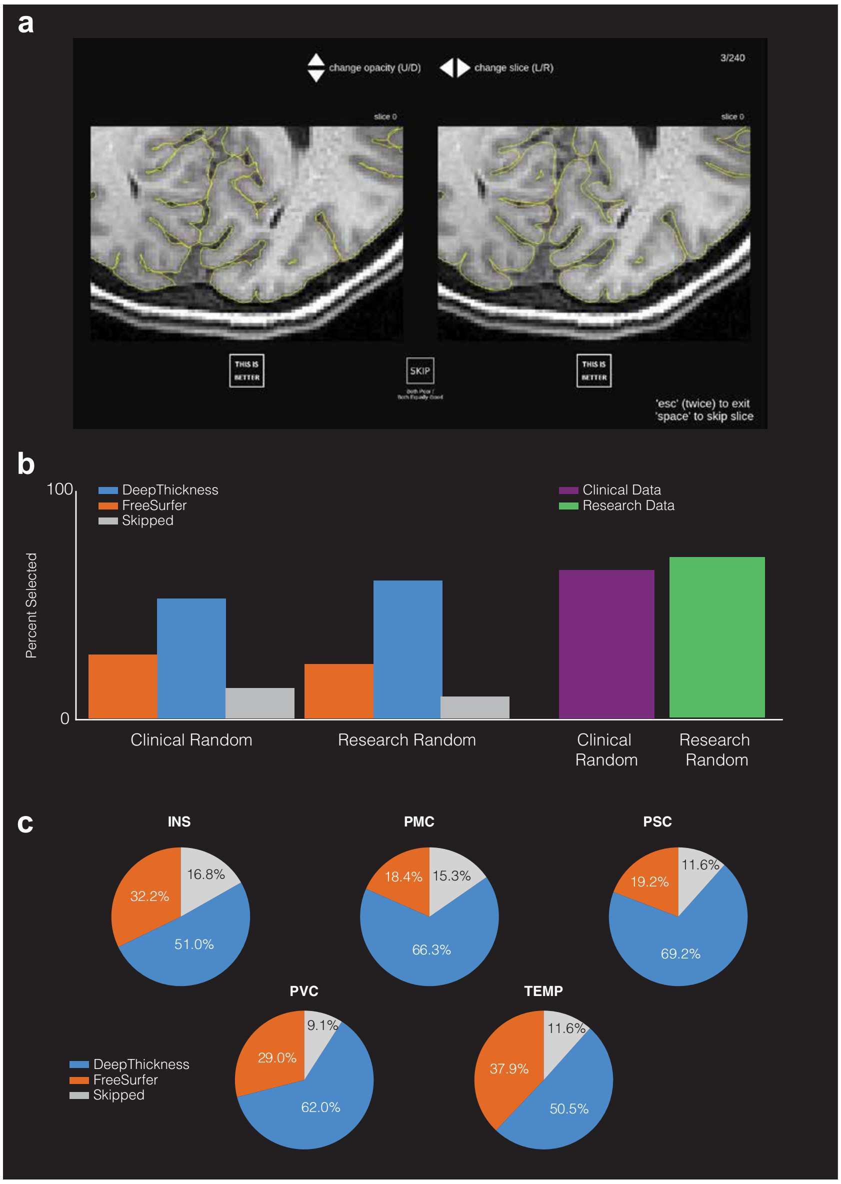


**Supplementary Fig 22**: **Expert Rater Results a** Example of behavioral evaluation trial in which neuroimaging experts assessed pial surface alignment to the T1w MRI by indicating their preference between two independently generated cortical surface reconstructions: one produced using FreeSurfer and the other using NeuroMorph. **b** Neuroexpert rater blind preference for FreeSurfer vs NeuroMorph surface alignment shown as a function of data provenance (left). Raters were instructed to skip if they believed both surfaces were both very poor/very good. Percentage of NeuroMorph preference split by data provenance (right). **c** Blind neuroimaging expert preference of surface in five regions of interest: Primary visual cortex, (V1 / Pericalcarine cortex), Primary motor cortex (Precentral gyrus), Primary somatosensory cortex (Postcentral gyrus), Temporal Lobe (Superior temporal gyrus), Insula Cortex.

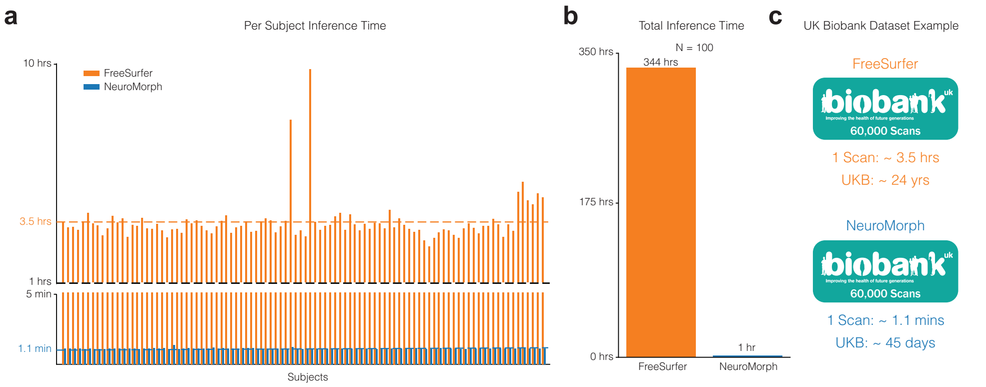


**Supplementary Fig 23**: **Inference Timing Comparison a** Inference time for each volume (n=100) using FreeSurfer's `recon-all` pipeline (orange) compared to NeuroMorph inference pipeline (blue). Dashed lines indicate mean inference time. **b** Total time to run all 100 volumes sequentially using 4 CPUs. **c** An applied example of approximate time required using FreeSurfer vs NeuroMorph to process the large UK Biobank dataset.

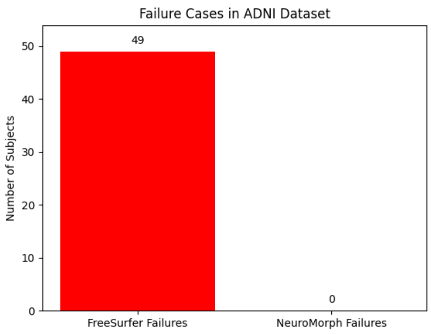


**Supplementary Fig 24**: **ADNI Failure Rates** The number of failures generated by FreeSurfer compared to NeuroMorph on the ADNI dataset (N=17329).
